# Biallelic variants in *DAP3* result in reduced assembly of the mitoribosomal small subunit with altered intrinsic and extrinsic apoptosis and a Perrault syndrome-spectrum phenotype

**DOI:** 10.1101/2024.08.19.24312079

**Authors:** Thomas B. Smith, Robert Kopajtich, Leigh A.M. Demain, Alessandro Rea, Huw B. Thomas, Manuel Schiff, Christian Beetz, Shelagh Joss, Gerard S. Conway, Anju Shukla, Mayuri Yeole, Periyasamy Radhakrishnan, Hatem Azzouz, Amel Ben Chehida, Monique Elmaleh-Bergès, Ruth I.C. Glasgow, Kyle Thompson, Monika Oláhová, Langping He, Emma M. Jenkinson, Amir Jahic, Inna A. Belyantseva, Melanie Barzik, Jill E. Urquhart, James O’ Sullivan, Simon G. Williams, Sanjeev S. Bhaskar, Samantha Carrera, Alexander J.M. Blakes, Siddharth Banka, Wyatt W. Yue, Jamie M. Ellingford, Henry Houlden, DDD Study, Kevin J. Munro, Thomas B. Friedman, Robert W. Taylor, Holger Prokisch, Raymond T. O’Keefe, William G. Newman

## Abstract

The mitoribosome synthesizes 13 protein subunits of the oxidative phosphorylation system encoded by the mitochondrial genome. The mitoribosome is composed of 12S rRNA, 16S rRNA and 82 mitoribosomal proteins encoded by nuclear genes. To date, variants in 12 genes encoding mitoribosomal proteins are associated with rare monogenic disorders, and frequently show combined oxidative phosphorylation deficiency. Here, we describe five unrelated individuals with biallelic variants in the *DAP3* nuclear gene encoding mitoribosomal small subunit 29 (MRPS29), with variable clinical presentations ranging from Perrault syndrome (sensorineural hearing loss and ovarian insufficiency) to an early childhood neurometabolic phenotype. Assessment of respiratory chain function and proteomic profiling of fibroblasts from affected individuals demonstrated reduced MRPS29 protein levels, and consequently decreased levels of additional protein components of the mitoribosomal small subunit, associated with a combined complex I and IV deficiency. Lentiviral transduction of fibroblasts from affected individuals with wild-type *DAP3* cDNA increased DAP3 mRNA expression, and partially rescued protein levels of MRPS7, MRPS9 and complex I and IV subunits, demonstrating the pathogenicity of the *DAP3* variants. Protein modelling suggested that *DAP3* disease-associated missense variants can impact ADP binding, and *in vitro* assays demonstrated *DAP3* variants can consequently reduce both intrinsic and extrinsic apoptotic sensitivity, DAP3 thermal stability and DAP3 GTPase activity. Our study presents genetic and functional evidence that biallelic variants in *DAP3* result in a multisystem disorder of combined oxidative phosphorylation deficiency with pleiotropic presentations, consistent with mitochondrial dysfunction.

## Introduction

Mitochondrial ribosomes (mitoribosomes) are present in the mitochondria of all eukaryotic cells. The function of the mitoribosome is to facilitate translation of mitochondrial mRNAs that exclusively encode components of the oxidative phosphorylation (OXPHOS) complexes. The mitoribosome consists of a small subunit (SSU) comprised of 30 mitoribosomal proteins (MRPs) and a 12S rRNA that binds mRNA and tRNA to ensure accurate initiation and decoding, and a large subunit (LSU) comprised of 52 MRPs, 16S rRNA, and mt-tRNA^Val^ that links a nascent polypeptide to the inner mitochondrial membrane via the OXA1L insertase ^1–4^. Formation of the mitoribosome is achieved through sequential steps. For the LSU, these steps can be divided into early, intermediate and late, whereas for the SSU these steps are only divided into early and late ^5^. Several human diseases are caused by germline variants in genes encoding mitoribosome proteins or assembly factors ^6^ (Table S1). Death-associated protein 3 (DAP3), also known as mitochondrial ribosomal small subunit 29 (MRPS29), is a GTP-binding protein of the mitoribosome SSU. The precise function of DAP3 within the mitoribosome remains unclear, but it is assembled into the SSU at an early stage, interacts extensively with the 12S rRNA and may associate with components of the inner mitochondrial membrane ^5,7^. DAP3 was initially identified as a pro-apoptotic protein ^8^ involved in interferon-γ-, tumor necrosis factor (TNF)-α- and FAS-induced cell death ^9^. DAP3 can also influence mitochondrial fission by modulating dynamin related protein phosphorylation, with DAP3 depletion resulting in decreased mitochondrial protein synthesis, ATP production and autophagy ^10^. Recently, DAP3 has also been linked to regulation of RNA editing and splicing in the context of cancer ^11,12^, further highlighting DAP3’s broad range of functions. To date, no *DAP3* variants have been reported in association with monogenic disorders.

Perrault syndrome is an ultra-rare, autosomal recessive condition characterized by sensorineural hearing loss (SNHL) in both sexes and primary ovarian insufficiency (POI) in 46, XX karyotype females (Pallister and Opitz, 1979). Neurological features are present in some affected individuals, often associated with brain white matter changes ^13^. As well as being clinically heterogeneous with variable degrees of severity, progression and age of onset of SNHL and POI in affected individuals ^14^, Perrault syndrome is remarkably genetically heterogeneous for such a rarely reported condition. To date, biallelic variants in eight genes are definitively associated with Perrault syndrome (Table S2). However, biallelic variants in other genes, including *RMND1*, *PEX6*, *MRPS7*, and *MRPL50* ^15–18^ have been identified in individuals with some features of Perrault syndrome, with a blended phenotype accounting for some diagnoses ^19^.

Despite this rich genetic architecture, potentially up to 50% of individuals with Perrault syndrome do not have a molecular diagnosis. Similarly, a large fraction of individuals with a suspected mitochondrial disease remain without a molecular diagnosis even after genome sequencing. Here, we present five individuals each with biallelic variants in *DAP3* (Table 1) with accompanying functional data providing evidence that *DAP3* variants result in decreased protein stability, reduced apoptotic sensitivity and impaired mitoribosomal assembly, leading to deficits consistent with mitochondrial disease. This study further underscores the importance of mitoribosome proteins in auditory and ovarian function.

**Table 1:**
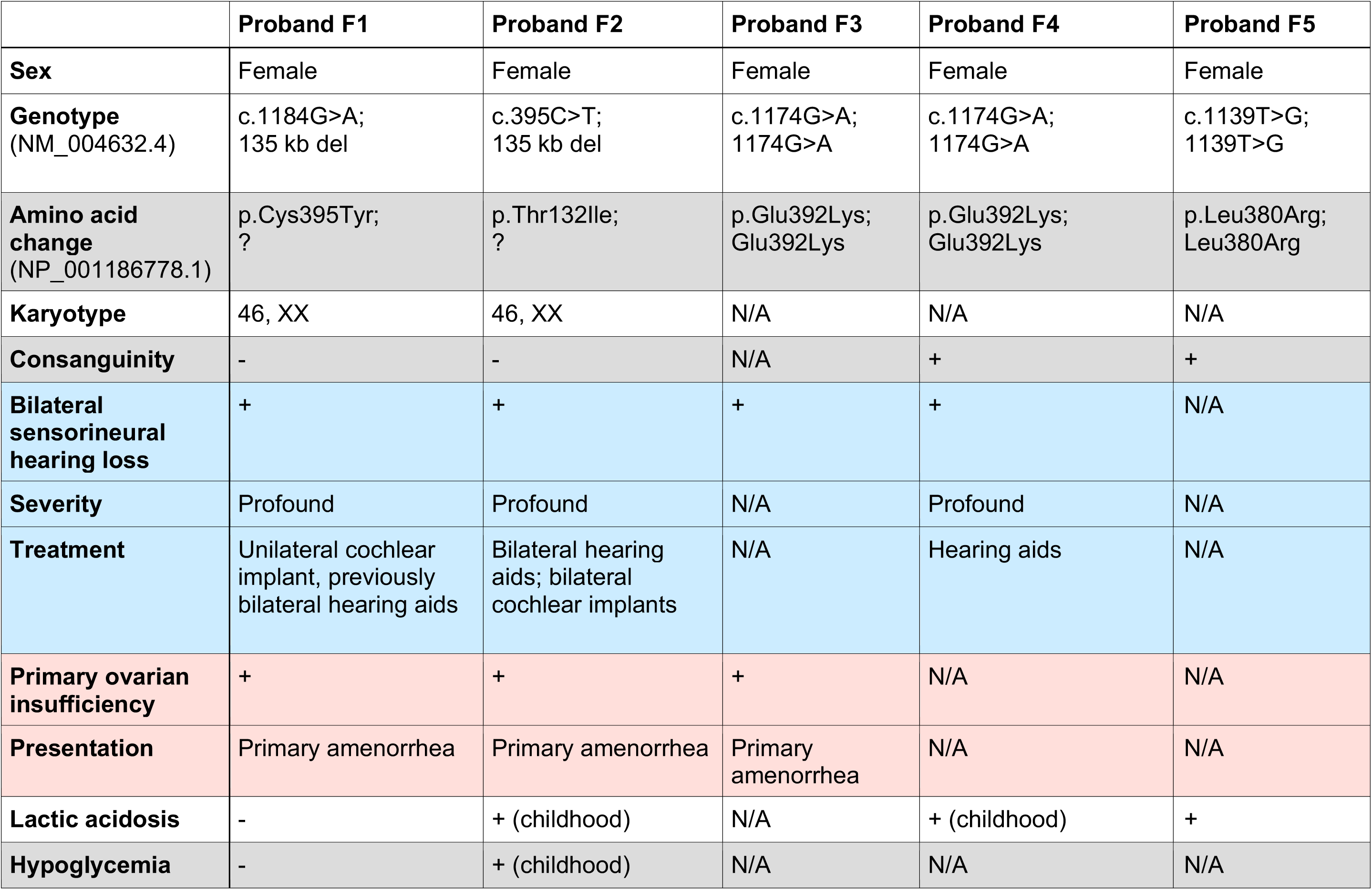
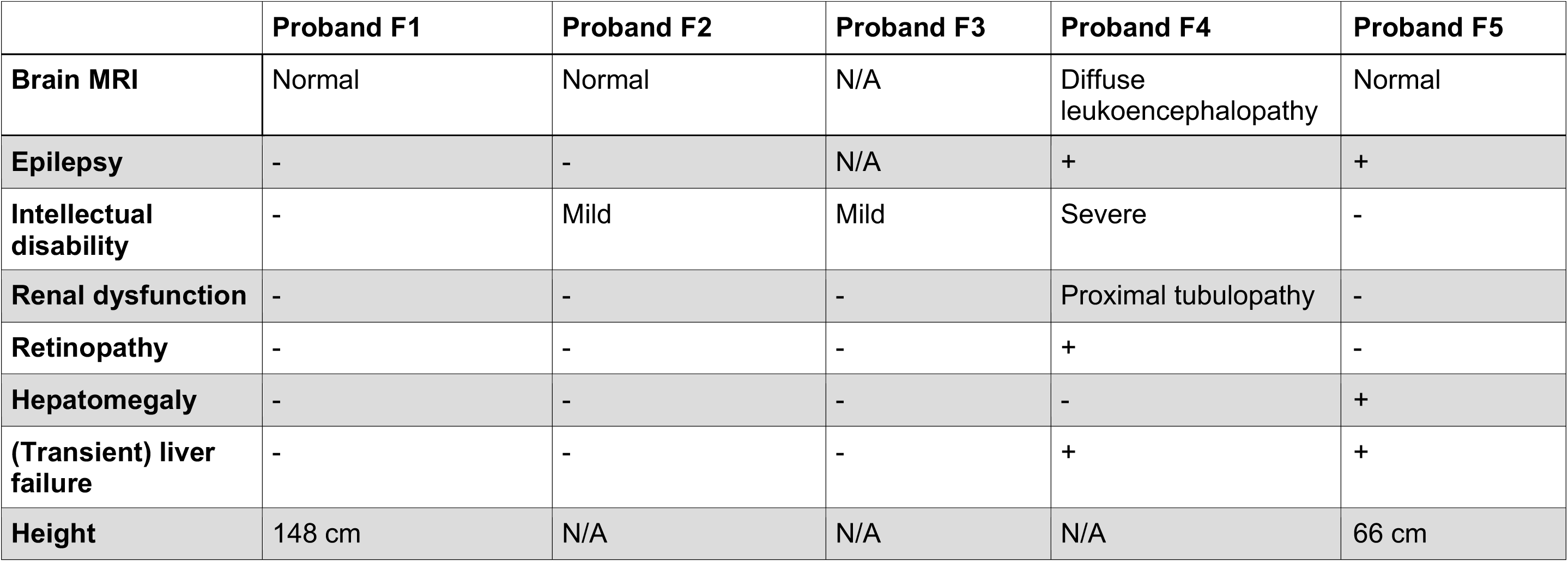
Phenotypic summary of individuals with *DAP3* variants identified in this study. Blue – categories linked to SNHL, red – categories linked to POI. N/A = not available.

## Material and methods

### Recruitment of research subjects

Individuals with clinical features of Perrault syndrome were recruited from the UK, Tajikistan, Tunisia and India through GeneMatcher ^20^, the Deciphering Developmental Disorders (DDD) project ^21^ and Centogene (https://www.centogene.com/). Informed consent for DNA analysis was obtained from study participants according to local institutional ethics requirements. Individuals (and/or their legal guardians) recruited in this study gave informed consent for their participation. The individual research studies received ethical approval by the National Health Service Ethics Committee (16/WA/0017 and 10/H0305/83) and The University of Manchester.

### Whole exome sequencing

WES was performed on DNA extracted from lymphocytes from individual F1:II-1. The SureSelect Human All Exon V5 Panel (Agilent Technologies) was used for library preparation and sequencing was performed on the HiSeq 2500 (Illumina) as previously described ^22^. Exome data for affected individuals in families F2-4 were <u>g</u>enerated as previously described ^21,23,24^. For F5:II-1, the TWIST Human Core Exome Plus exome capture kit was used, with the Illumina platform utilized for sequencing.

### Identification, amplification and confirmation of the DAP3 fusion product

A 135 Kb deletion encompassing *DAP3* was identified using the ExomeDepth (v1.1.6) software package ^25,26^. Read depth was approximately 0.5 times the aggregated depth indicating a single allele deletion. The fusion product and breakpoint region were confirmed in the F1 proband by Sanger sequencing using ABI big Dye v3.1 (Thermo Fisher Scientific Inc, Waltham, MA, USA) sequencing technology. Primers (Table S3) were designed to target polymorphisms distinguishing the two segmental duplications where the deletion breakpoints were situated.

### Maintenance of human dermal fibroblasts

Fibroblasts were cultured in high glucose Dulbecco’s Modified Eagle’s Medium (Sigma) with 10% foetal bovine serum (Sigma) and 10 mL/L penicillin-streptomycin (Sigma), at 37°C / 5% CO_2_.

### Fibroblast respiratory chain activity assays

Respiratory chain complex activities were assessed in fibroblasts from affected individuals F1 and F4, as outlined previously ^27^.

### RNA extraction, cDNA synthesis and RT-qPCR

Fibroblasts were seeded into 6-well plates (Corning) and were incubated at 37°C / 5% CO_2_ until approximately 90% confluent. Following one phosphate buffered saline (PBS) wash, RNA was extracted from cells using TRI-Reagent^®^ (Sigma), according to manufacturer’s instructions. Total RNA was converted to cDNA using the GoScript™ (Promega) Reverse Transcription System with random hexamers (Thermo Fisher) according to manufacturer’s instructions, normalizing all RNA concentrations to the lowest measured.

RT-qPCR reactions to assess 12S:16S ratios and mt-DNA gene expression were performed using 2 µM primer pairs, PowerUp™ SYBR™ Green Master Mix (Thermo Fisher) and 1 µL template cDNA. Primer sequences are listed in Table S3. The StepOnePlus Real-Time PCR System (Applied Biosystems) was used to measure fluorescence, using the Comparative CT reaction cycle programme. 2^-ΔΔCT^ values were calculated by the accompanying StepOnePlus v2.3 data analysis package, normalizing to *ACTB* (NM_001101.5) expression. 12S:16S ratios were calculated by totaling the 12S and 16S RQ values, then dividing the specific RQ value by the total value. All reactions were run in triplicate in 96-well plates. Data was presented using GraphPad Prism 9 throughout this study.

### Expression and purification of recombinant wild-type and variant *DAP3*

Purified DNA fragments comprised of truncated *DAP3* (DAP3Δ46) wild-type or disease-associated variants from amplified cDNA were inserted into the pMAL-c4X plasmid (New England Biolabs) at the multiple cloning site downstream of maltose binding protein (MBP), alongside a C-terminal 6x His-tag using NEBuilder® HiFi DNA Assembly Master Mix (New England Biolabs) according to the manufacturer’s instructions. A pMAL-c4x vector containing MBP fused to only the 6x His-tag was also produced for a negative control. All primer sequences for site-directed mutagenesis and mutagenic oligonucleotides are listed in Tables S3-S4. Following confirmation via Sanger sequencing (Eurofins Genomics), plasmids were transformed into Rosetta 2 (DE3) *E. coli* cells (Novagen) and cultured in Overnight Express TB medium (Novagen) at 19°C for 72 hours. Pellets were resuspended in lysis/wash buffer comprised of 20 mM Tris-Cl pH 7.4, 150 mM NaCl, 0.1 mM DTT, 20 mM imidazole (Sigma) and 15% glycerol. All purified proteins were captured and separated by affinity chromatography utilizing the 6x His-tag. His-tagged proteins were then eluted in lysis/wash buffer containing 250 mM imidazole. Selected fractions were then dialyzed overnight at 4°C in 20 mM Tris-HCl pH 8, 200 mM NaCl, 2 mM DTT and 15% glycerol. Proteins were then centrifuged at 17,000 x g for 10 minutes at 4°C, and the supernatants were frozen at -80°C.

### GTPase assays

GTPase assays were conducted using the GTPase-Glo™ Assay (Promega) in white opaque 96-well plates (Greiner Bio-One) in accordance with the manufacturer’s guidelines. A final concentration of 5 µM DAP3 protein, 5 µM GTP and 1 mM DTT was selected for use in the GTPase reaction, which ran for one hour at room temperature. Luminescence of residual GTP converted to ATP was measured using the BioTek Synergy H1 microplate reader (Agilent) 10 minutes after addition of detection buffer, with reactions conducted in duplicate over three independent assays. Residual GTP was calculated as a percentage using a no protein control, with an MBP-His protein control ran in parallel to ensure observed GTPase activity was DAP3-specific. Data was collected using Gen5 v2.07 software (Agilent).

### Proteomic analysis

Fibroblasts from F1:II-1 and F4:II-1 were processed and analyzed through an established proteomics pipeline to quantify the protein levels of both DAP3, and components of the mitoribosome and respiratory chain complexes. Two parameters of the protocol previously described ^28^ have been modified: Peptide fractionation was carried out using high pH reverse phase instead of trimodal mixed-mode chromatography and TMT-labeling was carried using TMT 11-plex instead of TMT 10-plex reagent. For data normalization, quantification and detection of aberrant protein expression, a denoising autoencoder based approach OUTRIDER2 was employed (termed PROTRIDER in Kopajtich *et al.* 2021).

### Apoptosis assays

Control and affected individual fibroblasts were seeded in opaque, white 96 well plates (Greiner Bio-One) at a density of 15,000 cells per well and incubated for 24 hours at 37°C / 5% CO_2_. Cells were treated with either 1 µM staurosporine (Cayman) for 4.5 hours to induce the intrinsic apoptotic pathway, 0.05 µg/mL or 0.5 µg/mL TNF-α (Sigma) in combination with 10 µg/mL cycloheximide (Cayman) for 24 hours to induce the extrinsic apoptosis pathway, or with suitable controls (0.01% DMSO and 10 µg/mL cycloheximide). Apoptotic activity was quantified using the Caspase-Glo® 3/7 Assay System (Promega), as per manufacturer’s instructions.

### Thermal shift assay (TSA)

TSA was performed using the Protein Thermal Shift^TM^ Dye Kit (Fisher Scientific) as per the manufacturer’s instructions in 96 well plates using the StepOnePlus Real-Time PCR System. 1 µg of recombinant MBP-DAP3 protein was subjected to melt-curve analysis in triplicate, progressing from 25°C to 90°C with a 1% temperature ramp rate. Melting temperature (T_m_) was derived by plotting melt curves of temperature against fluorescence intensity, selecting the temperature at which peak fluorescent intensity was detected.

### Lentiviral transduction of *DAP3* cDNA

A third-generation lentiviral construct was assembled using VectorBuilder, inserting full-length *DAP3* cDNA upstream of T2A:EGFP under the control of an EF1α short form (EFS) promoter. Following confirmatory Sanger sequencing and lentiviral packaging, fibroblasts from affected individuals and controls were seeded in 12-well plates at a density of 40,000 cells per well for RNA extraction, or into T25 flasks (Corning) at a density of 200,000 cells per flask for immunoblotting. Cells were immediately transduced in combination with 5 µg/mL Polybrene (Sigma), then incubated for 24 hours at 37°C and 5% CO_2_. Cells were washed three times with PBS, then growth media was replaced. After 72 hours post-transduction, cells were washed 3 times with PBS and processed as required. Subsequent RNA extraction, cDNA synthesis and qPCR analysis were conducted as described above.

### SDS-PAGE and immunoblotting

Cells were pelleted and lysed in 50 µl Pierce™ IP Lysis Buffer (Thermo Scientific) supplemented with 50x protease inhibitor cocktail (Promega) on ice, then agitated for 30 minutes at 4°C and centrifuged at 13,000 rpm for 15 minutes. Samples were mixed 1:1 with 2X SDS-PAGE sample buffer and heated to linearise protein, then ran on a 4-12% polyacrylamide gel made in-house at 180V for 60 minutes alongside the Precision Plus Protein Dual Color Standards (Bio-Rad) ladder. Proteins were transferred onto a 0.45μm PVDF blotting membrane (GE Healthcare) using a Trans-Blot Semi-Dry Transfer Cell System (Bio-Rad) for 30 minutes at 20V. The membrane was washed with 1x TBS-Tween and blocked with 5% milk for 1 hour with agitation. Primary antibodies specific to MRPS7 (Abcam, ab224442), MRPS9 (Abcam, ab187906), the five antibodies provided in the Total OXPHOS Human WB Antibody Cocktail (ATP5A, UQCRC2, SDHB, COXII, and NDUFB8) (Abcam, ab110411) and beta-actin (ProteinTech; 20536-1-AP, 66009-1-Ig) were incubated overnight at 4°C in block with agitation. Dilutions were 1:200 (MRPS7, MRPS9), 1:500 (Total OXPHOS) and 1:5000 (beta-actin) respectively. After washing, secondary antibodies were incubated with the membrane for 1 hour at 1:10,000, and were as follows: IRDye® 800CW Goat anti-Rabbit IgG (LI-COR, 926-32211) and IRDye® 680RD Goat anti-Mouse IgG antibody (LI-COR, 926-68070). Blots were washed in TBS-Tween and visualised with the LICOR Odyssey FC imaging system using the 600, 700 and 800 channels. Quantification was achieved using LICOR Image Studio and beta-actin was used to normalise band intensities.

### DAP3 localisation in mouse organ of Corti

The NIH Animal Care and Use Committee approved protocol #1263-15 to T.B.F. for mouse use. C57Bl/6J mice at postnatal day 3 (P3), P10, and P14 were euthanized, the cochleae were removed and fixed with 4% paraformaldehyde in PBS for 2 hours at room temperature (RT). The samples were microdissected, and the organ of Corti was permeabilized with 0.5% Triton-X100 in PBS for 30 minutes followed by three 10 minute washes with 1X PBS. Nonspecific binding sites were blocked with 5% normal goat serum and 2% BSA in PBS for 1 hour at RT. Samples were incubated for 2 hours at RT with primary antibodies (Table S5) in blocking solution, followed by several rinses with PBS. Then, samples were incubated with secondary antibodies (Table S5) for 30 minutes at RT, washed several times with PBS, mounted with ProLongGold Antifade staining reagent with DAPI (Molecular Probes, Invitrogen, Carlsbad, CA, USA) and examined using a LSM780 confocal microscope (Zeiss Microimaging Inc, Oberkochen, Germany) equipped with 63X, 1.4 N.A. objective.

### Mouse organ of Corti Helios Gene Gun transfection

Inner ear sensory epithelium cultures were prepared from P1 C57Bl/6J mouse organ of Corti and transfected with plasmid DNA using the gene gun method as previously described ^29^. Briefly, the organ of Corti spiral was dissected in Leibowitz cell culture medium (Invitrogen, Carlsbad, CA, USA) and attached to a glass-bottom Petri dish (MatTek, Ashland, MA, USA), coated with rat tail collagen and maintained at 37°C and 5% CO_2_ in DMEM supplemented with 7% FBS for 1–3 days. Cultures were transfected using a Helios gene gun (Bio-Rad, Hercules, CA, USA). Gold particles of 1.0 μm diameter (Bio-Rad, Hercules, CA, USA) were coated with DAP3-EGFP plasmid DNA at a ratio of 2 μg of plasmid DNA to 1 mg of gold particles and precipitated onto Tefzel tubing, which was cut into individual cartridges containing approximately 1 μg of plasmid DNA. Samples were bombarded with gold particles from one cartridge per culture by using 110-120 psi of helium. After an additional 8 hours to 4 days in culture, samples were fixed in 4% paraformaldehyde and stained using the same method as the tissue samples above.

### Statistical analysis

Statistical analyses were accomplished using GraphPad Prism 9 (GraphPad) software, performing one-way or two-way ANOVAs using either Dunnett’s or Tukey’s multiple comparisons tests where appropriate, as indicated in the figure legends. Statistical significance was defined as a *p*-value < 0.05.

## Results

Full phenotypic details are available from the authors on request.

Family F1 is a non-consanguineous white European family (Figure 1A) with an affected female proband who was diagnosed with bilateral, profound SNHL in early childhood (Figure S1A). She presented with primary amenorrhea leading to a diagnosis of Perrault syndrome. Otherwise, she had normal development and intellect. She had a successful unilateral cochlear implant as an adult. The mother is unaffected, whilst the father is deceased from an unrelated condition.

**Figure 1:**
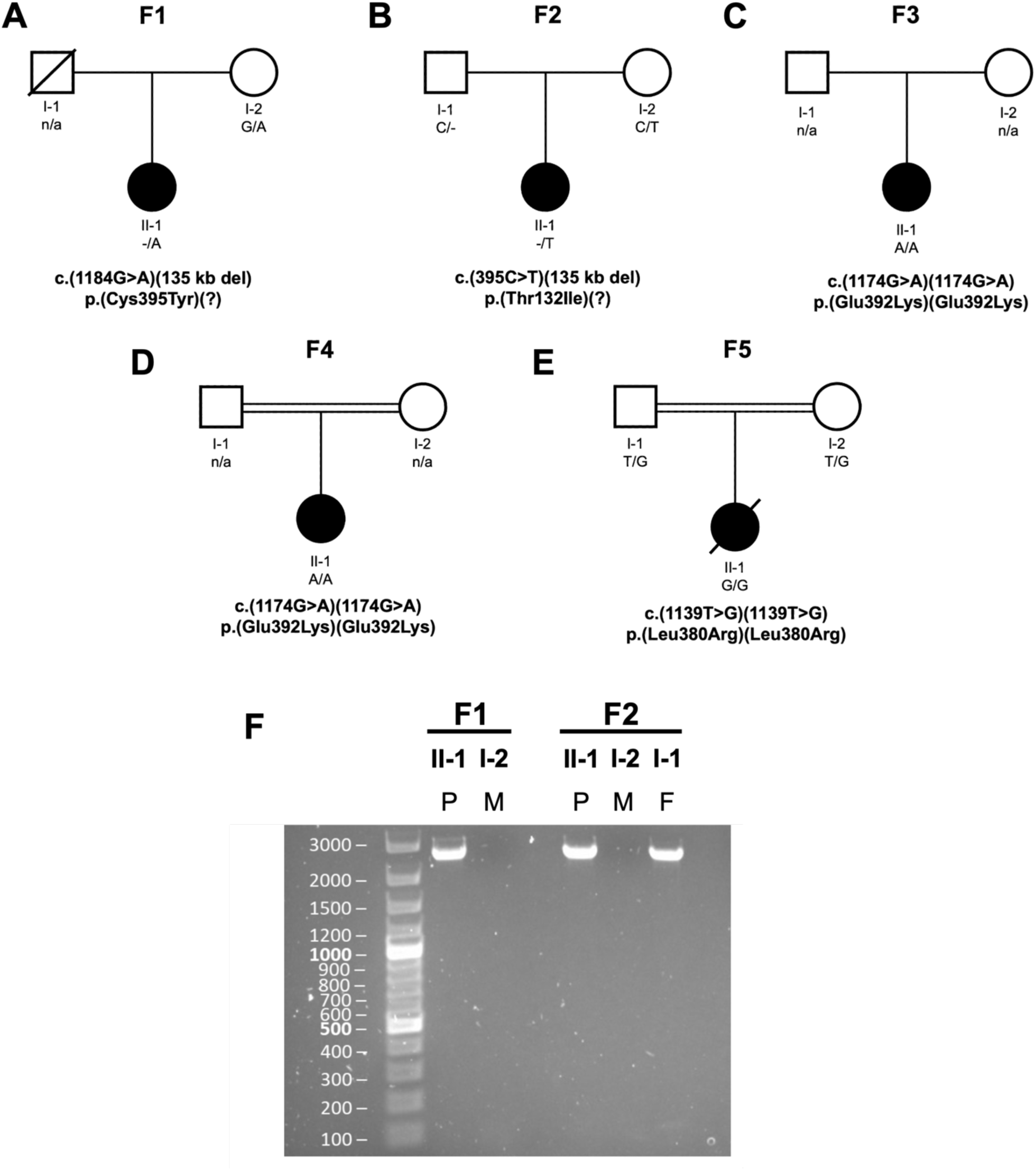
Family pedigrees and characterization of the *DAP3* deletion fusion product present in F1 and F2. (A-E) Pedigrees for the five families, with known segregation and variant details listed. All variants are annotated against the *DAP3* reference sequence NM_004632.4. (F) PCR analysis of F1 and F2 DNA using gel electrophoresis to detect a fusion product for the 135 kb deletion. P = proband, M = mother, F = father.

WES initially uncovered no putative pathogenic variants in known Perrault syndrome genes, but additional filtering revealed the F1 proband was compound heterozygous for the missense variant *DAP3* (NM_004632.4:c.1184G>A, p.(Cys395Tyr)), *in trans* to a 135 kb deletion identified with multiplex ligation-dependent probe amplification (MPLA).

Breakpoints were established to recombine between Chr1(GRCh38):g.155641696-155777755, which encompasses *DAP3* as well as *YY1AP1*, *GON4L* and *MSTO2P*. WES data revealed no rare variants in these other genes. The *DAP3* variant p.(Cys395Tyr) was confirmed as heterozygous in the unaffected mother by Sanger sequencing, however it is unknown whether the deletion was a *de novo* event or inherited paternally.

Family F2 is a non-consanguineous white European family (Figure 1B), ascertained through the Deciphering Developmental Disorders (DDD) study ^21^. The proband was diagnosed with bilateral SNHL in early childhood. As an adult she had bilateral cochlear implants (Figure S1B). She presented with primary amenorrhea (Figure S1C). In early childhood, she experienced recurrent episodes of ketosis, lactic acidosis and hypoglycemia and has mild intellectual disability. As an adult her brain MRI was normal.

Trio WES data in F2 identified a maternally inherited *DAP3* (NM_004632.4:c.395C>T p.(Thr132Ile)) missense variant *in trans* to a paternally inherited 135 kb deletion. A PCR fusion product of the same size as in the F1 proband was detected in the F2 proband, and her unaffected father (Figure 1F). The repetitive nature of this chromosomal region made it impossible to confirm whether the breakpoints are identical in both families.

Family F3 was identified through Centogene. The proband is a young woman from Central Asia (Figure 1C). She presented with bilateral SNHL of unknown severity, primary amenorrhea, mild intellectual disability and developmental delay. No further clinical information is available for this family. WES revealed the proband was homozygous for the *DAP3* (NM_004632.4: c.1174G>A, p.(Glu392Lys)) missense variant.

Family F4 is a consanguineous family of North African ancestry (Figure 1D). The affected proband is a girl who presented in early childhood with neurological impairment following a febrile infection with seizures. Brain MRI revealed diffuse leukoencephalopathy, with a lactate peak on spectroscopy (Figure S2). She had profound SNHL, transient liver failure and proximal tubulopathy. Electroretinogram studies revealed retinopathy. CSF and blood lactate levels were 4.5 mmol/L and 5-7 mmol/L respectively (normal ranges 1.1 – 2.4 and ≤ 2mmol/L), with an increased lactate/pyruvate ratio. Respiratory chain analysis activity testing on muscle cells revealed a complex IV deficiency, with borderline complex I deficiency. She has severe intellectual disability. The proband was homozygous for the *DAP3* (NM_004632.4:c.1174G>A, p.(Glu392Lys)) missense variant.

Finally, family F5 is a consanguineous family from the Indian sub-continent with a family history of neonatal and infant mortality (Figure 1E). The affected individual presented in early childhood with fever, vomiting and lethargy. Further testing revealed hepatosplenomegaly and lactic acidemia. Brain MRI was unremarkable. No hearing evaluation was completed. The provisional diagnosis was mitochondrial disorder with hepatic failure, and she died shortly after presentation. WES revealed the proband was homozygous for the *DAP3* (NM_004632.4:c.1139T>G, p.(Leu380Arg)) missense variant.

All affected DAP3 residues are well-conserved; representing 65% (p.Thr132Ile), 91% (p.Leu380Arg), 66% (p.Glu392Lys), and 73% (p.Cys395Tyr) of the respective amino acid positions across 300 orthologs using Consurf (Figure 2A). All substituted amino acids are also not present in any orthologs ^30^. Multiple *in silico* analyses predict these variants to be pathogenic or deleterious (Table S6). The four missense variants are either absent or have extremely low allele frequencies in the gnomAD v4.0 dataset (Table S7) ^31^, in further support of pathogenicity.

**Figure 2:**
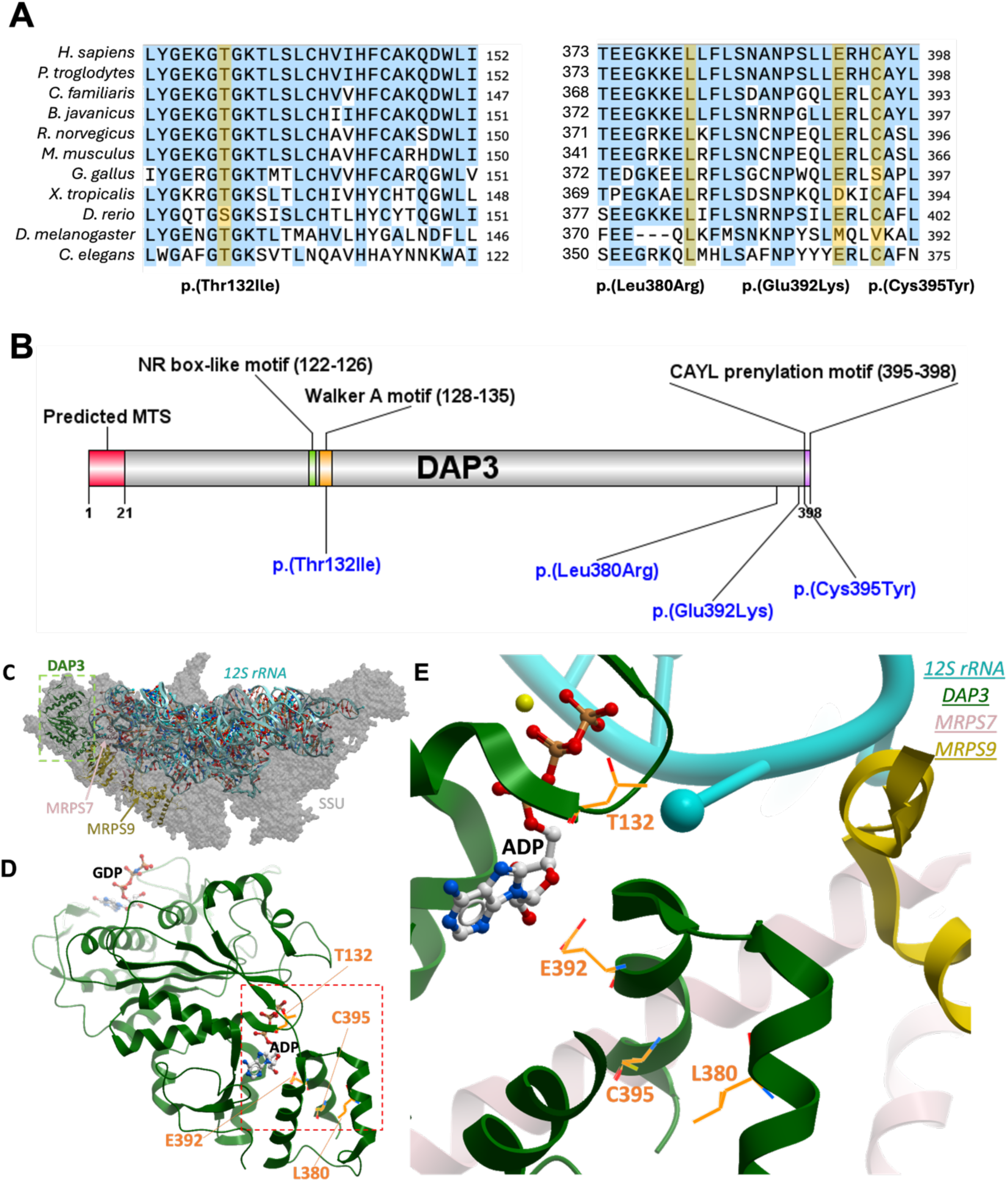
DAP3 variant residue conservation status, variant locations, and structural context. (A) Evolutionary conservation of affected DAP3 residues, with a broad selection of species highlighted. Variant amino acids highlighted in black, with yellow signifying matching to the associated human residue. Sequences aligned using Jalview 2.11.2.7 ^32^. The DAP3 reference sequence used for these species are listed accordingly: *H. sapiens*: NP_001186778.1; *P. troglodytes*: XP_016802675.2; *C. familiaris*: XP_038527847.1; *B. taurus*: NP_001106765.1; *R. norvegicus*: NP_001011950.2; *M. musculus*: NP_001158005.1; *G. gallus*: XP_040546712.1; *X. tropicalis*: NP_001016002.1; *D. rerio*: NP_001092207.1; *D. melanogaster*: NP_523811.1; *C. elegans*: AAD20727.1. (B) Overview of *DAP3* variant locations, with additional regions or domains of interest for additional context. MTS – mitochondrial targeting sequence, NR – nuclear receptor, CAYL – cysteine alanine tyrosine leucine (final 4 residues at the DAP3 C-terminus). (C) Cryo-EM structure of human mitochondrial ribosome small subunit at 2.40 Å resolution (PDB id: 7P2E), highlighting DAP3 (green), MRPS7 (rose) and MRPS9 (yellow) subunits. (D) Cartoon representation of DAP3 bound with GDP and ADP. (E) ADP binding site of DAP3 in proximity to the four sites of mutation (orange sticks).

We next inspected the site of variants at the protein level (Figure 2B), based on the recently determined structure of human mitoribosome SSU ^33^. DAP3/MRPS29 is localized in the head region of SSU (Figure 2C), close to the interface with the LSU. Three affected residues sit around a nucleotide binding site, currently believed to bind ATP (Figure 2D). Threonine 132 sits within a Walker A motif (**G**EKGT_132_**GKT**), which is commonly associated with ATP or GTP/GDP binding ^34^. Cysteine 395 is located within a putative prenylation site (CAYL) at the DAP3 C-terminus ^35^ and is close to the interface with MRPS7, another mitoribosomal protein in which pathogenic variants have been associated with primary ovarian insufficiency ^36,37^ (Figure 2E). Glutamic acid 392 is located upstream of this prenylation site and is predicted to interact directly with ATP ^36^. Leucine 380 localizes in an α-helix that packs against MRPS7 and MRPS9.

To gain a deeper insight into the role of DAP3 in the inner ear, we used immunofluorescence to assess DAP3 localization within the mouse organ of Corti. Endogenous DAP3 was identified within murine organ of Corti but was irregularly distributed in hair cells before and after the onset of hearing, with higher expression observed in likely damaged cells, sometimes with misshapen nuclei (Figure S3A). Exogenous DAP3 tagged with EGFP was then transfected into the mouse organ of Corti and vestibular sensory epithelium using a Helios gene gun, to test how overexpression affected the inner ear sensory hair cells. Overexpression instigated co-localization of DAP3-EGFP with TOM20 in hair cells and diffuse staining within the cell body (Figure S3B), however, there was no discernible increase in cell death following DAP3 overexpression, indicating compensatory mechanisms may prevent unwarranted alterations to mitoribosomal and apoptotic functions in the inner ear. We also immunostained transfected inner ear epithelial explants with DAP3 antibodies and showed that antibody signal was increased in transfected cells only (Figure S3C), while remained practically undetectable in non-transfected cells, indicating the specificity of the antibody to DAP3 protein while pointing to very low levels of DAP3 in wild-type hair cells under normal conditions.

To investigate the pathogenicity of the *DAP3* variants, we characterized dermal fibroblasts obtained from the affected individuals in families F1 and F4. We assessed the respiratory chain complex activities of these fibroblasts in comparison to eight healthy control fibroblasts (Figure 3A). Interestingly, F1:II-1 proband fibroblasts exhibited a mild mitochondrial respiratory chain defect, with a clear decline in complex IV activity compared to control reference ranges. F4:II-1 fibroblasts exhibited a heightened respiratory chain defect, with a reduction in both complex I and complex IV activities (Tables S8-9). The F4 CI:CII ratio of activities was also decreased, indicative of a generalized disorder of mitochondrial translation.

**Figure 3:**
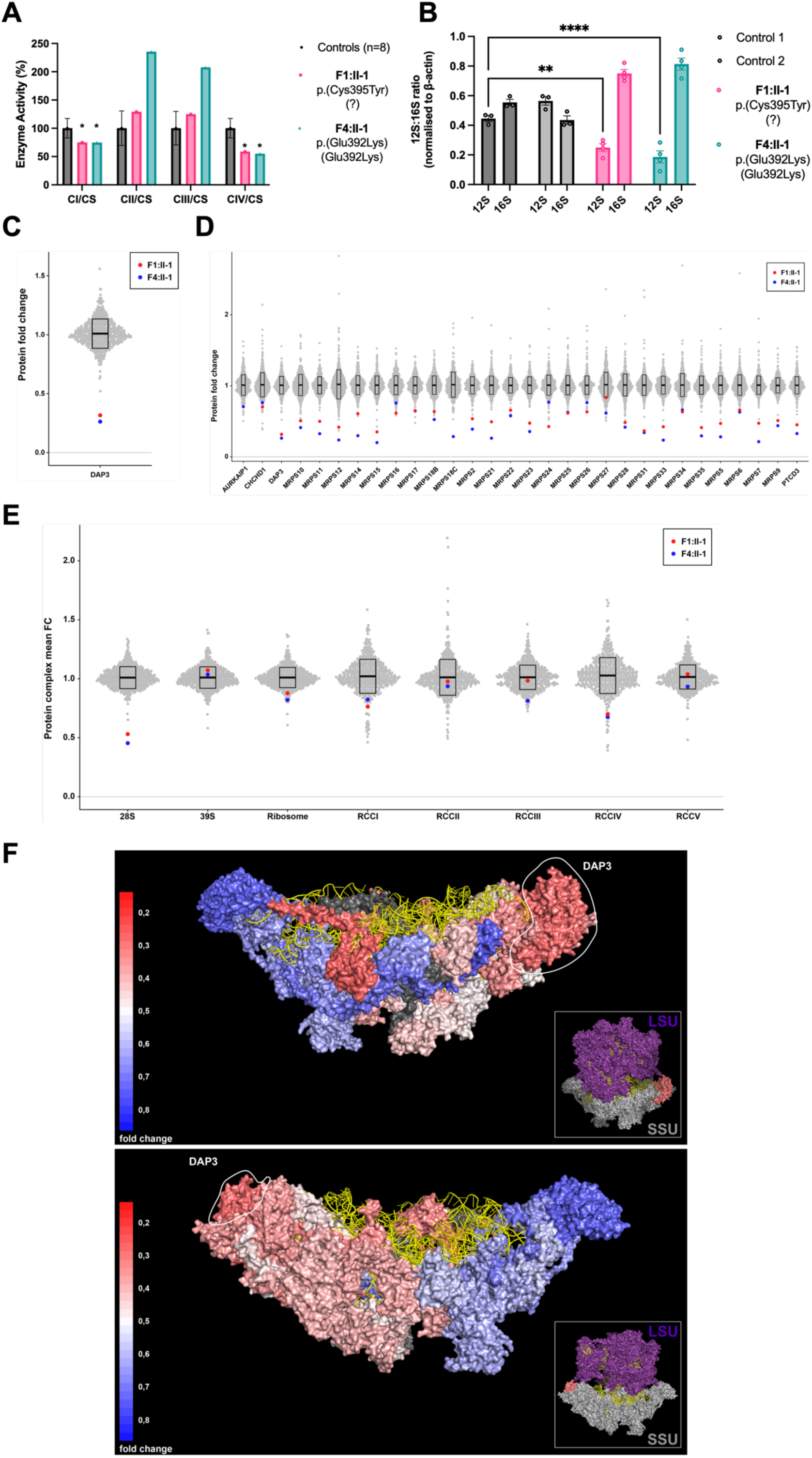
Functional and proteomic analysis of F1 and F4 proband fibroblasts reveal *DAP3* variants induce mitochondrial respiratory chain defects and decreased expression levels of small mitoribosomal subunit and OXPHOS components. (A) Mitochondrial respiratory chain enzyme activities in control (black), F1:II-1 (pink) and F4:II-1 (blue) fibroblast samples. Mean enzyme activities in control fibroblasts (n = 8) are set at 100%. Error bars represent standard deviation between the controls. * indicates enzyme activity is beyond control standard deviation values. (B) *MT-RNR1* (12S) and *MT-RNR2* (16S) expression levels in fibroblast cDNA. Data expressed as a ratio using relative quantification (RQ) values. Error bars represent the SEM. N = 3-4, **p < 0.01, ****p < 0.0001, two-way ANOVA with Tukey’s multiple comparisons test, comparing 12S RQ value of controls to affected individuals. (C) DAP3 protein levels in affected individual fibroblasts expressed as protein fold change compared to the mean of 512 fibroblast samples. (D) Protein fold change of all components of the mitoribosomal SSU in affected individual fibroblasts, compared to 512 controls. (E) Grouped mean fold change of all proteins comprising mitoribosome subunits, whole mitoribosome, and OXPHOS components compared to the mean values of 512 controls. (F) Cryo-EM structure (PDB id: 6VLZ) of mitoribosomal SSU with individual subunits colored according to their mean fold change values (of individuals F1:II-1 and F4:II-1) compared to the mean of 512 controls. Colors ranging from weakly reduced (blue) to strongly reduced (red) and two subunits (MRPS18C and MRPS38) in dark grey as no mean fold change values could be calculated. 12S ribosomal RNA is colored in yellow, DAP3 is marked by a circle, and the small inset shows the relative position within the 55S ribosome.

As DAP3 is a component of the small mitoribosomal subunit, we assessed if expression of *MT-RNR1* and *MT-RNR2*, which encode 12S and 16S rRNA respectively, was altered in F1:II-1 and F4:II-1 fibroblasts compared to healthy controls. The subsequent 12S:16S ratios were calculated using *MT-RNR1* and *MT-RNR2* relative quantification values to highlight specific contextual alterations to the small mitoribosomal subunit. The 12S component of the 12S:16S ratio was significantly reduced in F1:II-1 and F4:II-1 cDNA compared to controls, with the DAP3 p.Glu392Lys variant producing the strongest effect on *MT-RNR1* expression (p = <0.0001 (p.Glu392Lys;Glu392Lys) and 0.0019 (p.Cys395Tyr;?)) (Figure 3B). These data indicate an impairment of mitoribosomal assembly.

To assess whether *DAP3* variants influence levels of DAP3, mitoribosomal subunits, or other mitochondrial proteins, fibroblasts from F1:II-1 and F4:II-1 underwent proteomic analysis. The data were compared to a cohort of 512 individuals to visualize outliers. Interestingly, DAP3 was reduced to approximately 25% of mean levels in fibroblasts from both affected individuals, whilst also displaying the lowest DAP3 levels compared to any other individual in the dataset (Figure 3C). There was a remarkably consistent decrease in levels across all proteins constituting the small mitoribosomal subunit complex (Figure 3D) in fibroblasts from both affected individuals compared to the cohort, unless the protein was undetected in the mass spectrometry analysis (Table S10). When summing up the SSU and LSU overall, the two individuals with *DAP3* variants show the lowest SSU levels across the full cohort of samples, whilst the levels of LSU were not affected (Figure 3E).

The analysis of mitochondrial respiratory chain complexes revealed a reduction of complex I and complex IV subunits in both affected individuals. Moreover, F4:II-1 also displayed a reduction in complex III. This reduction agrees with the enzymatic analysis and reflect the downstream consequences on the translation of mtDNA-encoded respiratory chain complex subunits. To visualize subunit protein abundance in the context of its 3D structure, the data were mapped onto the Cryo-EM structure of the SSU (Figure 3F). Generally, proteins situated near DAP3 in the SSU are less abundant, with subcomplex formation more likely if situated on the opposite side to DAP3. These findings demonstrate independent evidence that *DAP3* variants impair assembly of the mitoribosomal SSU, impacting mitochondrial translation.

To assess whether disease-associated variants affect apoptosis, we cultured fibroblasts from F1:II-1 and F4:II-1 and challenged them with common effectors of intrinsic and extrinsic apoptosis pathways. We measured caspase-3 and caspase-7 activities with a commercial luminescence-based assay. Treatment with both staurosporine and TNF-α + cycloheximide significantly reduced caspase-3/7 release in affected individual fibroblasts compared to controls (Figure 4A). The fibroblasts from F4:II-1 exhibited a stronger apoptotic defect when challenged with intrinsic stimuli compared to the fibroblasts from F1:II-1. However, there were no significant differences between fibroblasts from affected individuals when treated with extrinsic stimuli.

**Figure 4:**
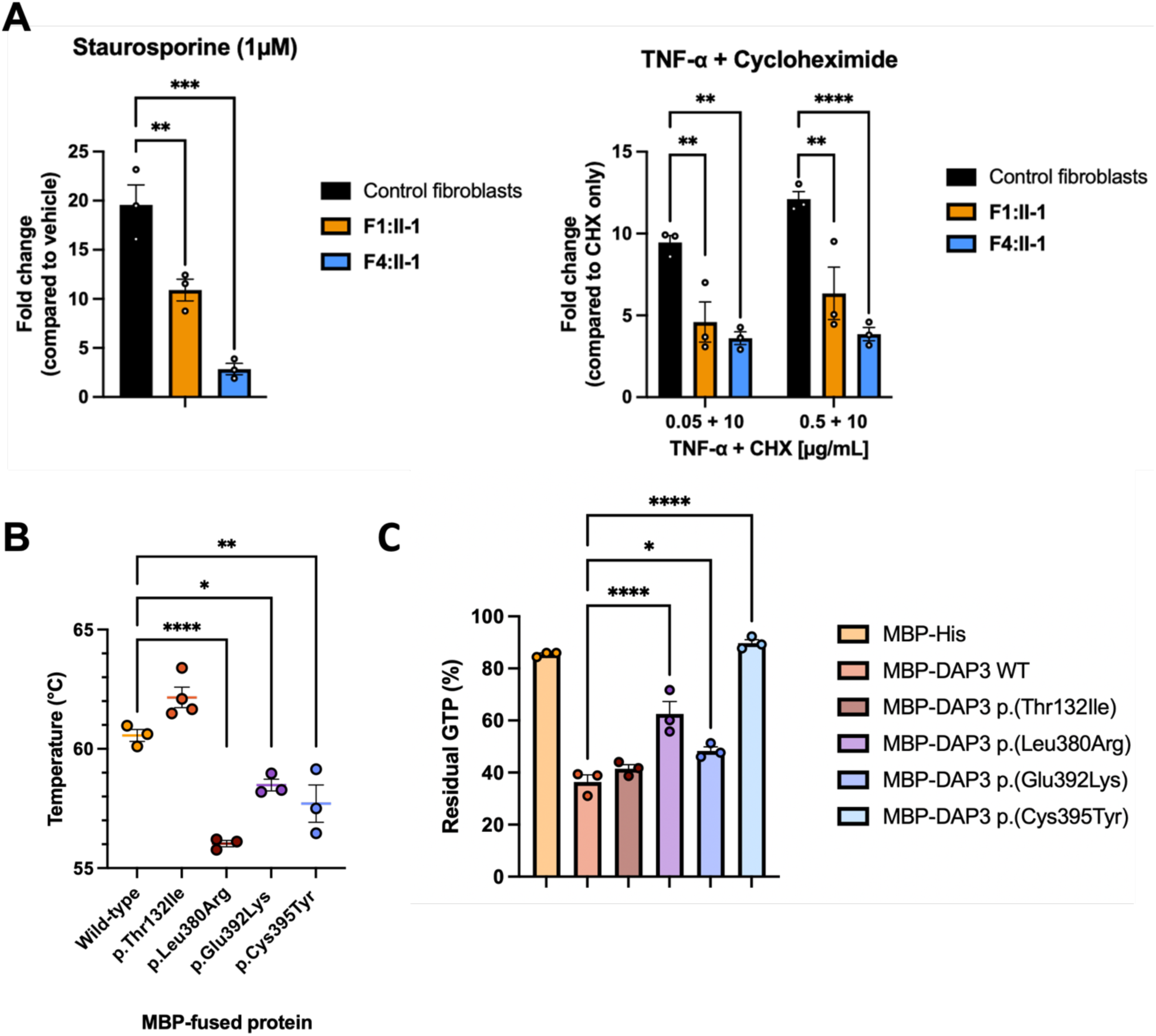
Functional analysis of both affected individual fibroblasts and recombinant DAP3 protein establish *DAP3* variants can diminish apoptotic sensitivity and destabilize DAP3 protein structure, impacting GTPase activity. (A) Assessment of caspase-3/7 release following stimulation of intrinsic and extrinsic apoptotic pathways. Affected individual fibroblasts were challenged in duplicate with staurosporine for 4.5 hours, or TNF-α + cycloheximide (CHX) for 24 hours before addition of assay reagent. Data expressed as fold change in luminescence signal compared to DMSO-treated or CHX-treated fibroblasts, with error bars representing SEM. N = 3, **p < 0.01, ***p < 0.001, ****p < 0.0001, using one-way ANOVA with Dunnett’s multiple comparisons test (staurosporine) or two-way ANOVA with Dunnett’s multiple comparisons test (TNF-α), comparing affected individual fibroblasts to control. (B) Thermal stability of recombinant wild-type and variant MBP-DAP3 protein. Data points represent average T_m_ of triplicate reactions. Error bars represent SEM. N = 3-4, *p < 0.05, **p < 0.01, ****p < 0.0001, using one-way ANOVA with Dunnett’s multiple comparisons test, comparing wild-type to variants. (C) GTPase activity of recombinant wild-type and variant MBP-DAP3 protein. Data presented as mean luminescence produced by residual GTP, with error bars representing SEM. N = 3, *p < 0.05, ****p < 0.0001, one-way ANOVA with Dunnett’s multiple comparisons test, comparing wild-type protein activity to variants.

To investigate the effect of *DAP3* variants on protein stability, we generated recombinant wild-type and variant DAP3 fused to maltose-binding protein (MBP). Thermal shift assay (TSA) and subsequent melt-curve analysis highlighted a significant T_m_ decrease in p.Leu380Arg, p.Glu392Lys and p.Cys395Tyr protein compared to wild-type (Figure 4B), indicating unfolding at lower temperatures and consequently reduced stability. Proteomic dissection of mitoribosomes indicated that DAP3 is the only GTP-binding protein in the SSU, suggesting that it could initiate or play a key role in mitochondrial protein synthesis ^10,38^. We hypothesized that *DAP3* variants could impair intrinsic GTPase activity, as disease-associated variants can impair DAP3 stability. Firstly, wild-type MBP-DAP3 exhibited GTPase activity *in vitro*. This GTPase activity was found to be significantly reduced with the DAP3 p.Leu380Arg, p.Glu392Lys and p.Cys395Tyr variant protein (p = <0.0001, 0.0206 and <0.0001 respectively), correlating with the TSA data (Figure 4C). The impact of these variants was variable, with a modest increase in residual GTP observed with p.Glu392Lys compared to wild-type. However, the p.Cys395Tyr variant increased residual GTP to the level observed with the negative control MBP-His, indicating low GTPase activity. Interestingly, there was no significant change in GTPase activity or thermal stability with p.Thr132Ile variant protein. These data suggest DAP3 variants can reduce protein stability, subsequently impairing ligand binding and GTPase activity.

To further confirm *DAP3* variant pathogenicity and specificity of their effect, we transduced fibroblasts from F1:II-1 and F4:II-1 with a lentiviral vector expressing wild-type *DAP3* to assess whether the mitoribosomal deficit could be rescued. *DAP3* mRNA expression increased in transduced cells, as expected (Figure 5A). Basal MRPS7 and MRPS9 levels were reduced in affected individuals, concordant with proteomic analysis. Following lentiviral transduction, immunoblotting also revealed a partial rescue of MRPS7 and MRPS9 protein levels in affected individual fibroblasts (Figure 5B), as well as in components of respiratory chain complex I (NDUFB8) and IV (COX II) (Figure 5C), changes which were not observed in transduced control fibroblasts.

**Figure 5:**
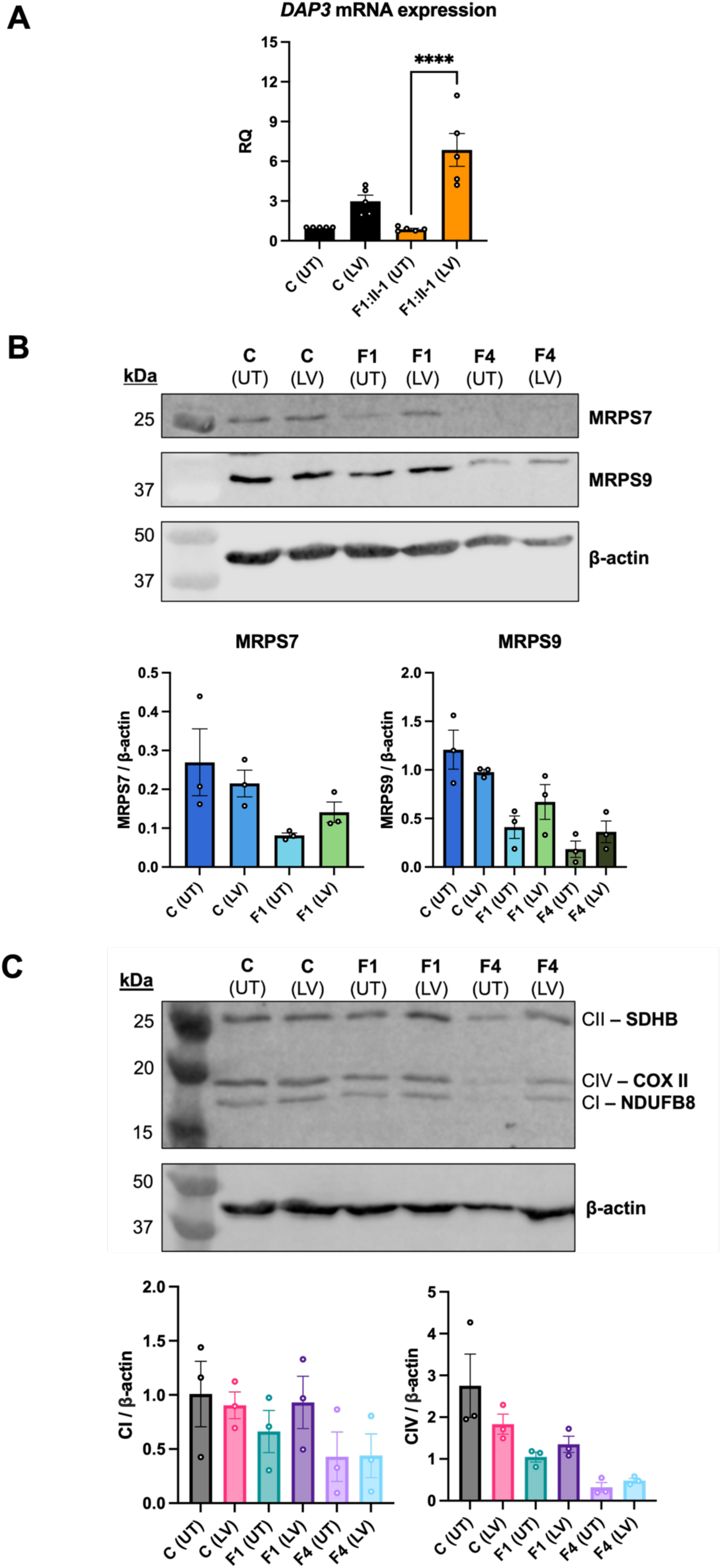
Lentiviral transduction of wild-type *DAP3* increases protein levels of MRPS7, MRPS9 and OXPHOS components in F1:II-1 and F4:II-1 fibroblasts. (A) Expression of *DAP3* mRNA in control and fibroblasts from F1:II-1 after lentiviral transduction (LV) of *DAP3* cDNA for 72 hours, or untransduced (UT). Each datapoint represents an averaged RQ value from triplicate reactions, using cDNA from independent transductions. Error bars represent SEM. N = 5, ****p < 0.0001, one-way ANOVA with Tukey’s multiple comparisons test. (B) Protein levels of MRPS7 and MRPS9 in control fibroblasts and fibroblasts from F1:II-1 and F4:II-1 after lentiviral transduction of *DAP3* cDNA for 72 hours. Beta-actin was used as a loading control and for densitometric analysis. Blots are representative of results from 3 independent biological repeats. MRPS7 levels were unable to be quantified in fibroblasts from F4:II-1. (C) Protein levels of SDHB, COX II and NDUFB8 in control fibroblasts and fibroblasts from F1:II-1 and F4:II-1 after lentiviral transduction of *DAP3* cDNA for 72 hours. Blots are representative of results from 3 independent biological repeats.

## Discussion

Using a range of genetic, molecular and proteomic techniques, this study reveals that biallelic *DAP3* variants are associated with a Perrault syndrome-spectrum phenotype. Most known Perrault syndrome-associated genes encode mitochondrial proteins with key roles in mitochondrial translation, which is consistent with DAP3 being a mitoribosomal SSU protein.

Phenotypes of individuals with *DAP3* disease-associated variants include a variety of features consistent with mitochondrial dysfunction, including lactic acidemia, neurological dysfunction, SNHL and POI, with variable expression (Table 1). Phenotypic severity ranges from classic Perrault syndrome, extending to childhood-onset neurological, developmental and multisystem abnormalities. The affected individuals homozygous for the same missense variant p.(Glu392Lys) have markedly different phenotypic presentations, with F4:II-1 affected with neurological, renal and retinal presentations. The probands from F1 and F2 have phenotypes which are less severe than in family F4. Both individuals from F1 and F2 have a hemizygous *DAP3* missense variant *in trans* to a 135 kb deletion, consistent with complete loss of function of one allele.

Diminished respiratory chain complex activities in fibroblasts from two affected individuals are consistent with a mitochondrial translation deficit (Figure 3A). Interestingly, the fibroblasts from F4:II-1 exhibited a more pronounced respiratory chain defect, with a clear reduction in both complex I and IV activities and diminished complex I:II ratios indicating a generalized disorder of mitochondrial translation. These data highlight that distinct *DAP3* variants have variable impacts on mitochondrial function.

The mtDNA-encoded 12S and 16S rRNA are essential components of the mitoribosomal SSU and LSU, respectively. They enable protein-RNA and protein-protein interactions which are key requirements for mitoribosome assembly and integrity ^39^. 12S:16S mRNA ratios have previously been evaluated to highlight specific discrepancies in mt-rRNA levels^40^. 12S rRNA is associated with Perrault syndrome due to disease-associated variants in the rRNA chaperone ERAL1, which also interacts with DAP3 ^40,41^. DAP3 is closely associated with the 12S rRNA, and when individual MRPS are diminished, 12S rRNA levels decline leading to SSU assembly defects ^17^. We hypothesized that 12S rRNA levels could be reduced in fibroblasts from affected individuals as DAP3 is assembled into the SSU at an early stage ^5^, and disrupted DAP3 function could lead to reduced mitoribosomal assembly and integrity. Indeed, 12S rRNA levels were significantly decreased in fibroblasts, whilst 16S rRNA levels were unchanged (Figure 3B).

Using sensitive quantitative proteomic profiling, biallelic *DAP3* variants were observed to confer a profile of mitochondrial ribosomal proteins typical for a SSU deficiency. Both fibroblasts from affected individuals demonstrated a clear, specific reduction in the levels of DAP3, but also all other SSU proteins, with all LSU proteins unaffected (Figure 3C-E). These data indicate that *DAP3* variants result in a specific impairment of SSU assembly. The loss of DAP3 could result in failure to assemble the mitoribosomal SSU triggering degradation of the 12S rRNA and other MRPs that require DAP3 or 12S rRNA as an assembly scaffold. The generalized decrease in SSU protein levels was more evident in fibroblasts from F4:II-1, consistent with her phenotypic severity. Proteomic profiles revealed the functional consequence of impaired assembly of mitoribosome as reduced mitochondrial translation of mtDNA encoded subunits of the respiratory chain complexes. Multiple respiratory chain complex proteins were reduced. Mainly complex I and IV mean protein abundance was affected in fibroblasts of both affected individuals, although complex III abundance was also reduced in F4:II-1, reflecting a more apparent generalized respiratory chain complex defect in this individual. The reduced respiratory chain complex activity of complex I and IV is consistent with other monogenic mitochondrial disorders ^42,43^, and variants in other MRPs, including *MRPS2, MRPS34* and *MRPL24* which result in impaired mitoribosome assembly ^44–46^. However, despite the common molecular effects, the clinical presentation of individuals with biallelic pathogenic variants in mitoribosomal proteins is heterogenous.

Interestingly, in the fibroblasts from F4:II-1, the largest reduction was observed in MRPS7 levels. Variants in *MRPS7* have been associated with clinical features overlapping Perrault syndrome ^17,37^. DAP3 and MRPS7 are predicted to interact extensively, including at Cys395 ^36^, which may explain the shared phenotypic spectrum. Variants in the gene encoding 12S rRNA are associated with sensorineural, non-syndromic deafness ^47^, suggesting altered MRPS7 and 12S rRNA interactions due to their reduced abundance may account for the SNHL in individuals with variants in *DAP3*.

We mapped protein mean fold change values seen with *DAP3* variants onto a Cryo-EM structure of the SSU to visualise SSU protein abundance within a structural context (Figure 3F). Interestingly, subunit abundance does not always reflect its proximity to DAP3. For example, MRPS12 and MRPS15 levels were substantially decreased in both sets of fibroblasts. Both MRPS12 and MRPS15 are assemble late to the SSU and are distant from DAP3, yet both interact extensively with 12S rRNA ^5,48^, which may reflect the importance of steady-state 12S levels for successful assembly and stability.

Intriguingly, four SSU proteins (MRPS7, MRPS12, MRPS15, MRPS33) exhibited marked depletion, especially in fibroblasts from F4:II-1. The 392 residue is predicted to interact with ATP, a ligand which stabilizes DAP3, and two neighboring residues Ser389 and Arg393 also contact an unpaired base of the 12S rRNA which may also stabilize the mitoribosome ^49^. This observation may indicate the DAP3 p.(Glu392Lys) variant is more likely to induce structural defects that impair initial sub-complex assembly, and reduction in SSU proteins in this individual. Taken together, these data demonstrate *DAP3* variants effect a global reduction in SSU protein levels leading to impaired mitoribosome assembly and mitochondrial translation.

Previous data has suggested multiple Perrault syndrome-associated genes are distinctly expressed within the spiral ganglion neurons of the cochlea, predicting variants could interfere with auditory signal transmission ^13^. Mouse organ of Corti immunostaining did not suggest any obvious DAP3 localization patterns to specific compartments of the inner ear (Figure S3A), in contrast to Perrault syndrome associated PRORP which was localized to synapses and nerve fibers of hair cells ^50^. Diffuse DAP3 cytoplasmic staining partially overlapping with mitochondrial marker TOM20 staining was observed before and after the onset of hearing in wild-type mice in some hair cells which sometimes appeared damaged with misshapen nuclei (Figure S3A). Exogenous overexpression shows increased mitochondrial localization in hair cells without cell damage (Figure S3B-C). These data imply DAP3 is present within the mouse inner ear at relatively low levels with no clear localization profile but might be upregulated in some stress conditions, indicating that SNHL in individuals with Perrault syndrome may have diverse gene specific etiologies.

Treating fibroblasts from affected individuals with intrinsic and extrinsic apoptosis mediators revealed a decrease in apoptotic sensitivity compared to controls (Figure 4A). These data contrast with previous studies evaluating the role of DAP3 in apoptosis, which have described variable effects on extrinsic receptor-mediated cell death but no desensitizing effects reported via the intrinsic mitochondrial-mediated death mechanism ^10,51^. It is possible that DAP3 disease-associated variants or the subsequent reduction in DAP3 abundance could affect interactions with known mediators of the intrinsic apoptosis pathway, or cells damaged by impaired mitoribosome assembly could induce non-specific mechanisms that impair the ability of the cell to detect or stimulate components of the intrinsic apoptosis pathway. DAP3 has been proposed to act as an adapter protein for death-inducing signaling complexes involved in the extrinsic pathway, recruiting FADD to TRAIL receptors (DR4 and DR5) in a GTP-dependent manner which may be aided by DAP3-binding protein death ligand signal enhancer (DELE1) ^52,53^. Diminished and unstable DAP3 protein can lead to reduced death receptor assembly, and subsequent signal transduction could explain the reduced sensitivity of fibroblasts from affected individuals to TNF-α.

Melt-curve analysis revealed *DAP3* variants p.Leu380Arg, p.Glu392Lys and p.Cys395Tyr exhibited significantly lower melting temperatures than wild-type, demonstrating these C-terminal variants destabilize DAP3 (Figure 4B). These data broadly correlate with the GTPase results, indicating the decreased stability could impair ligand binding and indirectly interfere with subcomplex assembly and mitochondrial protein synthesis. The DAP3 p.Leu380Arg variant conferred the most severe effect on thermal stability, consistent with the severe clinical phenotype. The p.Thr132Ile variant had no effect on thermal stability, so how this variant results in disease remains undetermined. However, residue 132 sits within the highly conserved Walker A motif [GxxxxGK(S/T)] which is necessary for ATP binding ^34^.

Proteins with GTPase activity can act as molecular switches and regulate a series of cell signaling events, including mitoribosome assembly. Mitoribosome assembly GTPases, such as ERAL1 and GTP-binding protein 10 (GTPBP10), can participate as rRNA chaperones and assembly factors, as well as conducting rRNA modifications and subunit quality control ^54,55^. DAP3 is the only predicted GTPase of the mitoribosome ^38^, but the functional extent of its putative GTPase activity is unclear. A recent structural study suggested that DAP3 GTPase activity is independent of the translation cycle of the mitoribosome. However, GDP binding to DAP3 was predicted to be required for efficient mitochondrial protein synthesis via enhanced stability of the DAP3 β-hairpin at residues 208-216 ^49^, highlighting the importance of DAP3 GDP binding to global mitoribosome function. We sought to understand if recombinant DAP3 exhibited intrinsic GTPase activity, and if GTPase activity was affected by the disease-associated variants. DAP3 variants p.Leu380Arg, p.Glu392Lys and p.Cys395Tyr significantly reduced GTPase activity, but DAP3 p.Thr132Ile had no effect (Figure 4C). These DAP3 residues are not located close to the GDP binding region, which suggested reduced stability and improper folding may non-specifically destabilize the GDP binding pocket. Residual GTPase activity does not appear to correlate with phenotype severity, as the individual who is compound heterozygous for p.Cys395Tyr has the least severe clinical presentation. Specific variants such as DAP3 p.Cys395Tyr may also alter key DAP3 modifications such as farnesylation of the CAYL motif ^35^. However, it is unclear if DAP3 is sufficiently prenylated *in vivo* for this modification to contribute to phenotypic variability ^38^.

Rescue experiments were performed to further verify *DAP3* variant pathogenicity. Lentiviral transduction of wild-type *DAP3* increased *DAP3* mRNA expression (Figure 5A). Immunoblotting revealed transduction increased MRPS7 levels in fibroblasts from F1:II-1, and MRPS9 levels in fibroblasts from F1:II-1 and F4:II-1 compared to untransduced cells, but not to control fibroblast protein levels (Figure 5B). This trend was not observed in control fibroblasts. The levels of CI and CIV subunits, NDUFB8 and COXII, respectively were also partially rescued in transduced fibroblasts, particularly in F1:II-1 fibroblasts (Figure 5C). This indicates that a partial rescue of depleted mitoribosomal SSU proteins in affected individual fibroblasts may aid stability of the mitoribosomal SSU, thus partially restoring CI and CIV biogenesis. This effect has been observed in several functional studies confirming variant pathogenicity in other SSU-encoding genes, further confirming that mitoribosome destabilization is associated with various heterogenous mitochondrial disorders ^45,46,56,57^. Collectively, these data indicate biallelic *DAP3* variants result in a Perrault syndrome spectrum phenotype, destabilizing the mitoribosome and impairment of mitochondrial translation.

Applying the ClinGen scoring criteria for gene-disease validity, we calculated a disease association score of 11 consistent with moderate evidence for disease association, which cannot be strengthened further without identification and characterization of additional affected individuals ^58^. However, the combined genetic, clinical and functional evidence outlined in this study provide confidence that biallelic *DAP3* variants are responsible for the described clinical presentations.

In summary, we have identified five independent families with biallelic variants in *DAP3* with a pleiotropic Perrault syndrome-associated phenotype, expanding the genetic heterogeneity of Perrault syndrome and further emphasizing the importance of mitochondrial translation in health and disease.

## Data Availability

All data produced in the present work are contained in the manuscript and supplementary information.

https://www.ncbi.nlm.nih.gov/clinvar

## Acknowledgements

We thank the families for their participation. This study was supported by the Medical Research Council (MR/W019027/1 ROK, RWT and WGN), Action on Hearing Loss (S35 Newman); Royal National Institute for the Deaf and Masonic Charitable Foundation (S60_Newman); Action Medical Research (GN2494); NIHR Manchester Biomedical Research Centre ((IS-BRC-1215-20007 and NIHR203308); Infertility Research Trust; Wellcome Trust ISSF pump-prime award [097820/Z/11/B]; the Wellcome Trust Centre for Mitochondrial Research (203105/Z/16/Z to RWT); the UK NHS Highly Specialised “Rare Mitochondrial Disorders of Adults and Children” Service (RWT); and The Lily Foundation (RWT) through PhD studentship funding to RICG; DBT/Wellcome Trust India Alliance for the study, “Centre for Rare Disease Diagnosis, Research and Training” (IA/CRC/20/1/600002); in part by the Intramural Research Program of the NIDCD at the NIH (DC000039 to TBF). The DDD study presents independent research commissioned by the Health Innovation Challenge Fund [grant number HICF-1009-003], a parallel funding partnership between Wellcome and the Department of Health, and the Wellcome Sanger Institute [grant number WT098051]. The research team acknowledges the support of the National Institute for Health Research, through the Comprehensive Clinical Research Network. The views expressed in the paper are those of the authors and not necessarily those of the funders.

## Ethical approval

All individuals or their guardians provided written informed consent in accordance with local regulations. Ethical approval for this study was granted by the National Health Service Ethics Committee (16/WA/0017) and University of Manchester. The NIH Animal Use Committee approved protocol 1263-15 to T.B.F. for murine studies.

## Author Contributions

TS, RK, LAMD, AS, HBT, CB, KT, MO, RICG, EMJ, AJ, IAB, MB, JEU, JOS, SGW, SSB, AJB, SC, JME generated laboratory data. MS, SJ, GSC, AS, MY, PR, HA, ABC, MEB, HH and WGN contributed genetic and clinical data. TBS, RK, LAMD, CB, SB, WWY, KM, TBF, RWT, HP, ROK and WGN designed and supervised the experiments and analyzed the data. TBS, ROK, WGN drafted the paper. All authors reviewed and critically contributed to the paper.

## Web Resources

dbSNP, https://www.ncbi.nlm.nih.gov/projects/SNP/

Ensembl Variant Effect Predictor (VEP), https://www.ensembl.org/info/docs/tools/vep/index.html

Exome Variant Server, http://evs.gs.washington.edu/EVS/

GenBank. https://www.ncbi.nlm.nih.gov/genbank/

GeneMatcher, https://genematcher.org/

GTEx, https://gtexportal.org/home/

gnomAD, http://gnomad.broadinstitute.org/

FoldX, http://foldxsuite.crg.eu/

LOVD, https://www.lovd.nl/

OMIM, https://www.omim.org/

MutationTaster, http://www.mutationtaster.org/

PolyPhen-2, http://genetics.bwh.harvard.edu/pph2/

SIFT, http://sift.bii.a-star.edu.sg

## Data and code availability

The *DAP3* variants were submitted to ClinVar (https://www.ncbi.nlm.nih.gov/clinvar/) (GenBank: NM_004632.4; accession numbers SCV004228990 – SCV004228993, VCV003066057.1).

