## Supplementary information for "Biallelic variants in *DAP3* result in reduced assembly of the mitoribosomal small subunit with altered intrinsic and extrinsic apoptosis and a Perrault syndrome-spectrum phenotype"

A

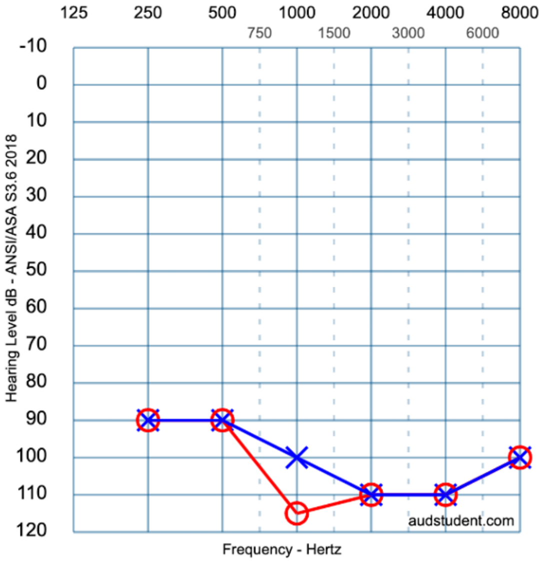


B

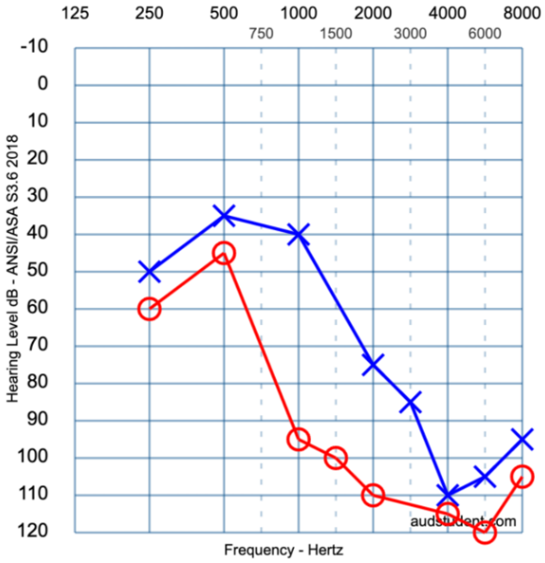


C

| Biochemical test | F2:II-1 (11-15 years) | Reference range (11-15 years, female) |
|---|---|---|
| Follicle Stimulating Hormone (FSH) | **101.5 IU/L** | < 0.1 - 12 IU/L |
| Luteinizing Hormone (LH) | **48.9 IU/L** | < 0.1 - 13.4 IU/L |
| Estradiol | **193.0 pmol/L** | < 20 - 87 pmol/L |

**Figure S1: Additional clinical information for the F1 and F2 probands.**

**(A)** Audiogram for individual F1:II-1. Hearing level in the left ear is represented by blue crosses, and the right ear by red circles. Designed using the AudGen online tool (version 0.6.3) (https://audsim.com/audgenJS/audgenjs.html).

**(B)** Audiogram for individual F2:II-1.

**(C)** Hormone profile for individual F2:II-1, consistent with hypergonadotropic hypogonadism.

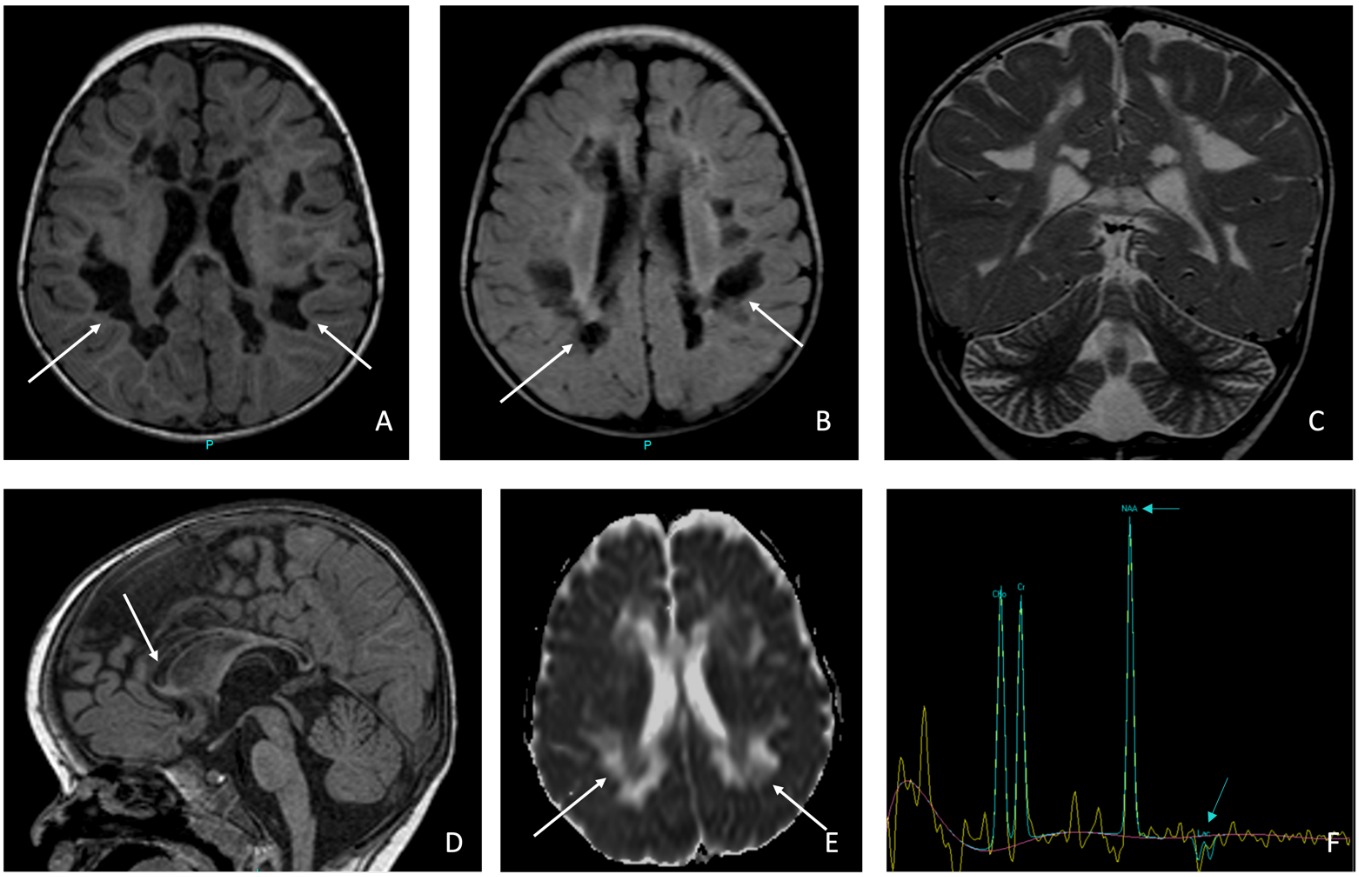


**Figure S2: Brain MRI and respiratory chain complex activities for affected individual F4:II-1.**

Individual F4:II-1, in early childhood: brain MRI shows a cavitating leukodystrophy.

**A-D** axial (A) and sagittal (D) T1-weighted images, **B** axial FLAIR at the same level than A, **C** coronal T2-weighted image through posterior part of the lateral ventricle and cerebellum, **E** axial diffusion-ADC map at the same level as A and B, **F** MR spectroscopy long TE monovoxel in the frontal white matter.

Bilateral T2/FLAIR white matter hyperintensity (B,C) with mostly symmetrical large areas of cavitation of the centrum ovale (A, B) sparing subcortical white matter, extending to the corpus callosum (D). Hemispheric cerebellar atrophy (C). No involvement of the basal ganglia, nor cerebellar peduncles and dentate nuclei (not shown). ADC map (E) shows increased diffusion of these areas without edges of restricted diffusion. MR spectroscopy (F) demonstrates a lactate peak (Lac) and a decreased NAA peak.

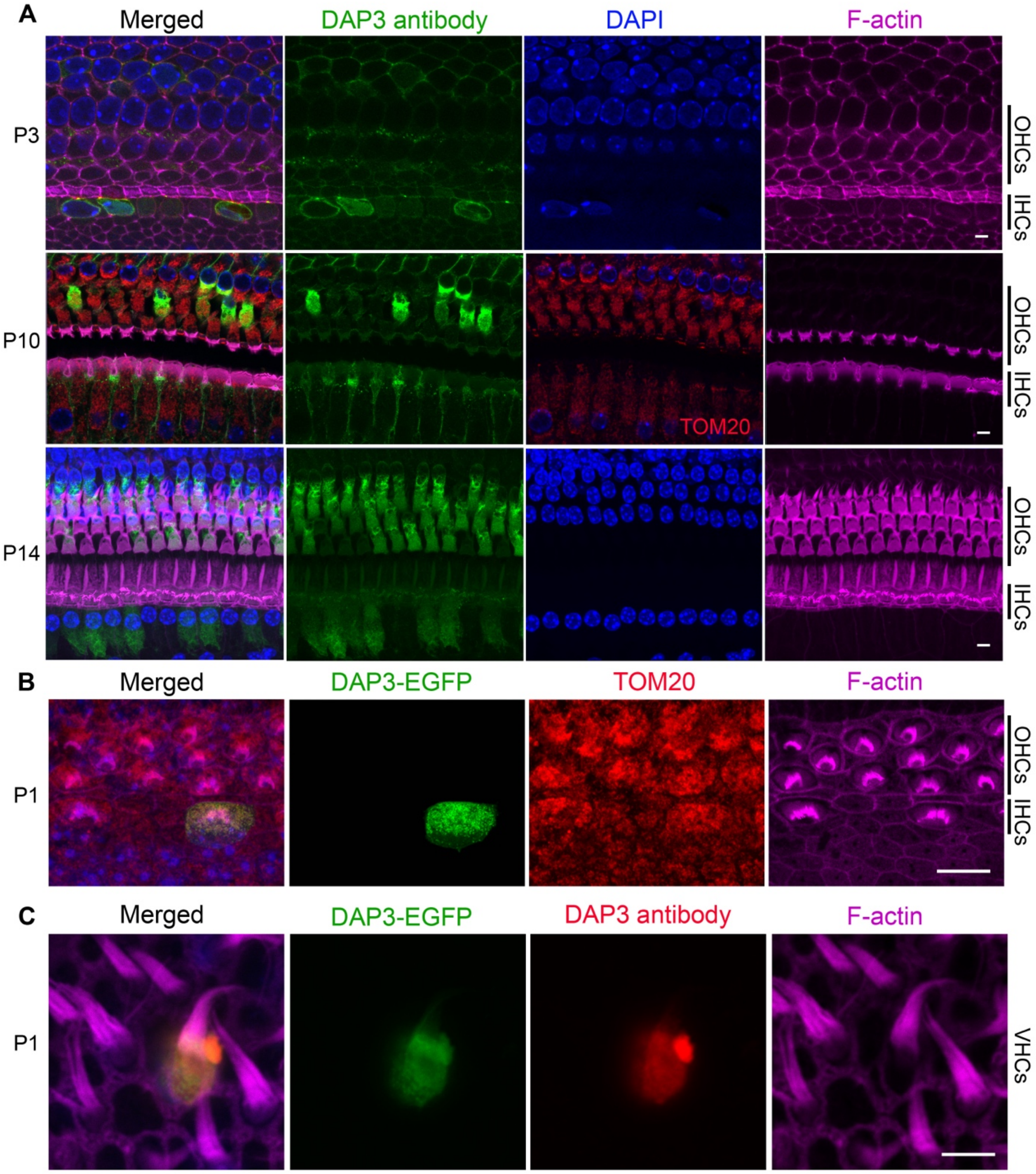


**Figure S3: DAP3 localization, antibody validation and exogenous overexpression in mouse inner ear sensory epithelium.**

**(A)** Confocal fluorescence microscopy images of whole mount organ of Corti samples from C57Bl/6J mice at postnatal days 3, 10 and 14 (P3, 10 and P14) showing localization of

DAP3 (green). Samples counterstained with the nuclear stain DAPI (blue) to visualize the nuclei of hair cells and rhodamine-phalloidin (magenta) to visualise the F-actin cytoskeleton and auditory hair cell stereocilia. P10 samples were also immunostained for the mitochondrial marker Tom 20 (red). Three rows of outer hair cells (OHCs) and one row of inner hair cell (IHCs) locations are indicated by black vertical bars on the right. Scale bars are 5 μm.

**(B)** A compressed Z-stack image of the organ of Corti explant culture from a P1 C57Bl/6J mouse transfected using Helios gene gun with the plasmid DAP3-EGFP (green). The sample was counterstained with TOM20 (mitochondrial marker, red), phalloidin (cytoskeletal marker, magenta) and a nuclear marker DAPI (blue); shown as merged and single channel images. Scale bar is 10 μm.

**(C)** Validation of DAP3 antibody (BD Biosciences). A hair cell from the inner ear vesibular epithelium of P1 C57Bl/6J mouse transfected with DAP3-EGFP (green). The sample is immunostained with DAP3 antibody (red, BD Biosciences) and counterstained by rhodamine-phalloidin (magenta) and DAPI (blue). Note that only transfected cell that overexpressed DAP3-GFP shows DAP3 antibody signal (red) indicating antibody's specificity. Scale bar is 5 μm.

| Gene (alternative name) | MIM Gene Reference | Phenotype | MIM Phenotype Reference | Multiple families/ variants | Reference |
|---|---|---|---|---|---|
| **EARLY-STAGE ASSEMBLY** | | | | | |
| **MRPS29** (DAP3) **(mS29)** | 602074 | SNHL, POI, hypoglycaemia, lactic acidemia | - | Multiple | This report |
| **MRPS7 (uS7m)** | 611974 | COXPD34: SNHL, renal and liver failure, lactic acidemia | 617872 | Multiple | 1 |
| **MRPS9 (uS9m)** | 611975 | NA | NA | NA | NA |
| **MRPS31** (IMOGN38) **(mS31)** | 611992 | NA | NA | NA | NA |
| **MRPS35 (mS35)** | 611995 | Failure to thrive, developmental/ intellectual delay, dysmorphism | NA | One | 2 |
| **MRPS39** (PTCD3) **(mS39)** | 619057 (614918) | COXPD51: Leigh syndrome, optic atrophy, SNHL | 619057 | One | 3 |
| **MRPS16** (RPMS16) **(bS16m)** | 609204 | COXPD2: agenesis of corpus callosum, dysmorphism, lactic acidemia | 610498 | One | 4 |
| **MRPS18B** (MRPS18-2) **(mS40)** | 611982 | NA | NA | NA | NA |
| **MRPS27** (KIAA0264) **(mS27)** | 611989 | Hereditary ataxia | NA | One | 5 |
| **MRPS34 (mS34)** | 611994 | COXPD32: Leigh syndrome, neurodevelopmental defects | 617664 | Multiple | 6 |
| **MRPS5 (uS5m)** | 611972 | NA | NA | NA | NA |

| | | | | | |
|---|---|---|---|---|---|
| **MRPS22** (C3orf5, RPMS22) **(mS22)** | 605810 | POI (type 7) COXPD5: Cardiomyopathy, dysmorphism, leukoencephalopathy, lactic acidemia | 618117 611719 | Multiple | 7,8 |
| **MRPS2 (uS2m)** | 611971 | COXPD36: SNHL, hypoglycemia, developmental delay, dysmorphism, lactic acidemia | 617950 | Multiple | 9 |
| **MRPS23 (mS23)** | 611985 | COXPD46: Liver disease | 618952 | One | 10 |
| **MRPS28 (bS1m)** | 611990 | COXPD47: failure to thrive, SNHL, liver impairment, dysmorphism hypoglycemia, lactic acidemia | 618958 | One | 11 |
| **MRPS12 (uS12m)** | 603021 | NA | NA | NA | NA |
| **MRPS17 (uS17m)** | 611980 | NA | NA | NA | NA |
| **MRPS11 (uS11m)** | 611977 | NA | NA | NA | NA |
| **LATE-STAGE ASSEMBLY** | | | | | |
| **MRPS6 (bS6m)** | 611973 | NA | NA | NA | NA |
| **MRPS38** (AURKAIP1) **(mS38)** | 609183 | NA | NA | NA | NA |
| **MRPS24 (uS3m)** | 611986 | NA | NA | NA | NA |
| **MRPS10 (uS10m)** | 611976 | NA | NA | NA | NA |
| **MRPS14 (uS14m)** | 611978 | COXPD38: Hypertrophic cardiomyopathy, lactic acidemia, dysmorphic features | 618378 | One | 12 |

| | | | | | |
|---|---|---|---|---|---|
| **MRPS33 (mS33)** | 611993 | NA | NA | NA | NA |
| **MRPS25 (mS25)** | 611987 | COXPD50: encephalopathy, microcephaly, short stature, dystonia | 619025 | Multiple | 13 |
| **MRPS26 (mS26)** | 611988 | NA | NA | NA | NA |
| **MRPS15 (uS15m)** | 611979 | NA | NA | NA | NA |
| **MRPS21 (bS21m)** | 611984 | NA | NA | NA | NA |
| **CURRENTLY UNCLEAR** | | | | | |
| **MRPS37** (CHCHD1) **(mS37)** | 608842 | NA | NA | NA | NA |
| **MRPS18C** (MRPS18-1) **(bS18m)** | 611983 | NA | NA | NA | NA |

**Table S1: Mitoribosomal SSU proteins grouped based on stage of assembly into the SSU, and their associated disorders.**

| Gene (definitive) | MIM Gene Reference | MIM Phenotype Reference | Putative protein function | MIM Genotype-phenotype relationship | Additional phenotypic observations | Reference |
|---|---|---|---|---|---|---|
| **HSD17B4** | 601860 | 233400, 261515 | Fatty acid beta-oxidation, steroid metabolism | Perrault syndrome, D-bifunctional protein deficiency | Leukodystrophy, ataxia | 14 |
| **HARS2** | 600783 | 614926 | Ligates histidine to tRNA in mitochondrial translation | Perrault syndrome | N | 15 |
| **LARS2** | 604544 | 615300, 617021 | Ligates leucine to tRNA in mitochondrial translation | Perrault syndrome, Hydrops, lactic acidosis, and sideroblastic anemia (HLASA) | Leukodystrophy, mitochondrial myopathy | 16 |
| **CLPP** | 601119 | 614129 | Proteolysis, mitoribosome formation and regulation | Perrault syndrome | Epilepsy, leukoencephalopathy | 17 |
| **TWNK** | 606075 | 616138, 271245, 609286 | Maintenance of mitochondrial DNA integrity | Perrault syndrome, mitochondrial DNA depletion syndrome 7, progressive external ophthalmoplegia with mitochondrial DNA deletions | Cerebellar atrophy, peripheral neuropathy | 18 |

| | | | | | | |
|---|---|---|---|---|---|---|
| **GGPS1** | 606982 | 619518 | Lipid synthesis, protein prenylation | Muscular dystrophy, congenital hearing loss, and ovarian insufficiency syndrome | Myopathy | 19 |
| **ERAL1** | 607435 | 617565 | Mitoribosome assembly | Perrault syndrome | N* | 20 |
| **PRORP** | 609947 | 619737 | Mitochondrial tRNA processing | Combined oxidative phosphorylation deficiency 54 (COXPD54) | White matter changes, developmental delay | 21 |

**Table S2: Genes definitively associated with Perrault syndrome.** The “additional phenotypic observations” column lists a brief selection of commonly observed symptoms that have been described alongside the classical Perrault syndrome phenotype.

** - Only one distinct variant has been published, so absence of neurological features or severe childhood multisystem phenotypes should be interpreted with caution until additional families have been identified.*

| Primer name | Sequence – 5′ to 3′ |
|---|---|
| **DAP3_BP-SeqInt_F** | TGGCAGAGTTAGCCGATGC |
| **DAP3_BP-SeqInt_R** | CCATTGGGAATGGATTTGACC |
| **12S/MT-RNR1_F** | TAGAGGAGCCTGTTCTGTAATCGAT |
| **12S/MT-RNR1_R** | CGACCCTTAAGTTTCATAAGGGCTA |
| **16S/MT-RNR1_F** | GCCTGCCCAGTGACACATG |
| **16S/MT-RNR1_R** | CACGGGCAGGTCAATTTCAC |
| **ActB_F** | GTGGATCAGCAAGCAGGAGT |
| **ActB_R** | GTAACAACGCATCTCATATTTGGAA |
| **DAP3_F** | CTGGCTGATACTACATATTCCAG |
| **DAP3_R** | GTTCAGGAAGCGCTCATTTGTA |
| **DAP3_380_F1_F** | TCGAGGGAAGGATTTCAATGGCTATTTCCCG |
| **DAP3_380_F1_R** | CAGCGAGGGGTTCGCGTTACTTAGGAACAG**C**CGCTC |
| **DAP3_380_F2_F** | GAAAAAAGAGC**G**GCTGTTCCTAAGTAACGCGAACCCCTCG |
| **DAP3_380_F2_R** | CTTGCCTGCAGGTCGACTGCCTGCAGGTCGACTCAAGAC |

**Table S3:** **Primer sequences used throughout this study. Red bold lettering indicates an altered base to reproduce *DAP3* variants identified in affected individuals.**

| Variant | Oligonucleotide sequence – 5′ to 3′ |
|---|---|
| c.395C>T p.(Thr132Ile) | **AAGACTTAGGGTTTTTCCTATTCCCTTCTCTCCATA** |
| c.1174G>A p.(Glu392Lys) | CGAATTCTTAGAGGTAGGCACAGTGCCGCT**TCAGCAGCGAGGGGTT** |
| c.1184G>A p.(Cys395Tyr) | CGAATTCTTAGAGGTAGGCA**TAGTGCCGCTCCAGC** |

**Table S4:** **Oligonucleotides designed to create DAP3 variant cDNA. Red bold lettering indicates an altered base.**

| Antibody target | Species raised in | Antibody conjugate | Supplier | Concentration |
|---|---|---|---|---|
| **DAP3** | Mouse | - | BDBiosciences | HeLa - 0.5μg/mL, Organ of Corti - 2.5μg/mL |
| **TOM20** | Rabbit | - | Santa Cruz | HeLa - 0.4μg/mL, Organ of Corti - 2μg/mL |
| **Cytoskeleton (F-actin)** | - | Alexa Fluor™ 647 | Thermo Fisher Scientific | HeLa - 1:50, Organ of Corti - 1:100 |
| **Mouse 2°** | Goat | Alexa Fluor™ 488 | Thermo Fisher Scientific | HeLa - 1:250, Organ of Corti - 1:400 |
| **Rabbit 2°** | Goat | Alexa Fluor™ 568 | Thermo Fisher Scientific | HeLa - 1:250, Organ of Corti - 1:400 |

**Table S5: Antibodies used for murine immunofluorescence studies.**

| | **c.395C>T p.(Thr132Ile)** | **c.1139T>G p.(Leu380Arg)** | **c.1174G>A p.(Glu392Lys)** | **c.1184G>A p.(Cys395Tyr)** |
|---|---|---|---|---|
| *Location* | Chr1(GRCh38):g.155725942-155725942C>T | Chr1(GRCh38):g. 155738184-155738184T>G | Chr1(GRCh38):g.155738219-155738219G>A | Chr1(GRCh38):g.155738229-155738229G>A |
| SIFT | **Deleterious** (0) | **Deleterious** (0) | **Deleterious** (0) | **Deleterious** (0) |
| Polyphen-2 (HumDiv) | **Benign** (0.315) | **Probably damaging** (1) | **Probably damaging** (0.976) | **Probably damaging** (0.937) |
| CADD | **Deleterious** (23.2) | **Deleterious** (28.8) | **Deleterious** (27.3) | **Deleterious** (25.2) |
| DANN (Rank score) | **Pathogenic** (0.676) | **Pathogenic** (0.812) | **Pathogenic** (0.979) | **Pathogenic** (0.798) |
| ClinPred | **Damaging** | **Damaging** | **Damaging** | **Damaging** |
| MutationTaster | **Disease causing** (0.999) | **Disease causing** (1) | **Disease causing** (1) | **Disease causing** (1) |
| REVEL | **Pathogenic** (0.549) | **Pathogenic** (0.865) | **Tolerable** (0.461) | **Pathogenic** (0.801) |
| AlphaMissense | **Ambiguous** (0.459) | **Likely pathogenic** (0.891) | **Likely pathogenic** (0.817) | **Likely pathogenic** (0.858) |

**Table S6: *In silico* pathogenicity predictions for *DAP3* variants identified in this study.**

| Variant | c.395C>T p.(Thr132Ile) | c.1139T>G p.(Leu380Arg) | c.1174G>A p.(Glu392Lys) | c.1184G>A p.(Cys395Tyr) |
|---|---|---|---|---|
| Allele count | **29** | **2** | **Absent** | **2** |
| Allele frequency | **0.000018** | **0.00000203** | - | **0.00000137** |
| Number of homozygotes | 0 | 0 | - | 0 |

**Table S7:** ***DAP3*** **variant allele frequencies in gnomAD v4.0.**

| Respiratory chain complex | F1:II-1 result | Reference ranges |
|---|---|---|
| Complex I / CS | **0.148** | 0.197 ± 0.034 |
| Complex II / CS | **0.283** | 0.219 ± 0.067 |
| Complex III / CS | **0.807** | 0.646 ± 0.192 |
| Complex IV / CS | **0.638** | 1.083 ± 0.186 |
| Complex I:II | **0.524** | 0.580 – 0.900 |

| Respiratory chain complex | F4:II-1 result | Reference ranges |
|---|---|---|
| Complex I / CS | **0.147** | 0.197 ± 0.034 |
| Complex II / CS | **0.516** | 0.219 ± 0.067 |
| Complex III / CS | **1.342** | 0.646 ± 0.192 |
| Complex IV / CS | **0.597** | 1.083 ± 0.186 |
| Complex I:II | **0.285** | 0.580 – 0.900 |

**Tables S8 & S9: Respiratory chain complex activities for both affected F1:II-1 (p.(Cys395Tyr)(?)) and F4:II-1 (p.(Glu392Lys)(Glu392Lys)) probands, with complex activities compared to citrate synthase (CS).**

| | | in pdb file 6VLZ | | P86317 | F4:II-1 | P149819 | F1:II-1 | mean | sd | |
|---|---|---|---|---|---|---|---|---|---|---|
| | | | | fold change | RGB-hex | fold change | RGB-hex | fold change | | RGB-hex |
| | | A | 12S-rRNA | | | | | | | |
| 0.150 | | d | MRPS7 | 0.213 | FF4343 | 0.473 | FFEDED | 0.343 | 0.184 | FF9898 |
| 0.175 | | F | MRPS12 | 0.237 | FF5252 | 0.418 | FFC9C9 | 0.327 | 0.128 | FF8D8D |
| 0.200 | | R | MRPS15 | 0.200 | FF3A3A | 0.351 | FF9D9D | 0.276 | 0.107 | FF6C6C |
| 0.225 | | X | MRPS33 | 0.235 | FF5151 | 0.423 | FFCDCD | 0.329 | 0.133 | FF8F8F |
| 0.250 | | O | DAP3 | 0.262 | FF6363 | 0.316 | FF8686 | 0.289 | 0.038 | FF7575 |
| 0.275 | | L | MRPS21 | 0.262 | FF6363 | 0.493 | FFFAFA | 0.378 | 0.163 | FFAFAF |
| 0.300 | | K | MRPS18C | 0.285 | FF7272 | #N/A | #N/A | #N/A | #N/A | #N/A |
| 0.325 | | Y | MRPS35 | 0.295 | FF7878 | 0.412 | FFC5C5 | 0.354 | 0.082 | FF9F9F |
| 0.350 | | c | MRPS5 | 0.281 | FF6F6F | 0.470 | FFEBEB | 0.376 | 0.133 | FFADAD |
| 0.375 | | G | MRPS14 | 0.297 | FF7A7A | 0.607 | B9B9FF | 0.452 | 0.219 | FFE0E0 |
| 0.400 | | b | PTCD3 | 0.328 | FF8E8E | 0.451 | FFDFDF | 0.389 | 0.087 | FFB6B6 |
| 0.425 | | W | MRPS31 | 0.342 | FF9797 | 0.366 | FFA7A7 | 0.354 | 0.017 | FF9F9F |
| 0.450 | | T | MRPS23 | 0.356 | FFA0A0 | 0.473 | FFEDED | 0.415 | 0.083 | FFC7C7 |
| 0.475 | | E | MRPS11 | 0.325 | FF8C8C | 0.500 | FFFFFF | 0.413 | 0.124 | FFC6C6 |
| 0.500 | | B | MRPS2 | 0.390 | FFB6B6 | 0.536 | E7E7FF | 0.463 | 0.103 | FFE7E7 |
| 0.525 | | N | MRPS28 | 0.418 | FFC9C9 | 0.483 | FFF4F4 | 0.450 | 0.046 | FFDEDE |
| 0.550 | | Q | MRPS10 | 0.412 | FFC5C5 | 0.507 | FAFAFF | 0.459 | 0.067 | FFE4E4 |
| 0.575 | | e | MRPS9 | 0.438 | FFD6D6 | 0.511 | F8F8FF | 0.474 | 0.051 | FFEEEE |
| 0.600 | | P | CHCHD1 | 0.763 | 5252FF | 0.702 | 7A7AFF | 0.733 | 0.043 | 6666FF |
| 0.625 | | S | MRPS22 | 0.578 | CCCCFF | 0.655 | 9999FF | 0.617 | 0.054 | B2B2FF |
| 0.650 | | J | MRPS18B | 0.525 | EFEFFF | 0.637 | A5A5FF | 0.581 | 0.079 | CACAFF |
| 0.675 | | a | AURKAIP1 | 0.707 | 7777FF | #N/A | #N/A | #N/A | #N/A | #N/A |
| 0.700 | | M | MRPS25 | 0.629 | ABABFF | 0.616 | B3B3FF | 0.622 | 0.009 | AFAFFF |
| 0.725 | | D | MRPS6 | 0.633 | A8A8FF | 0.660 | 9696FF | 0.646 | 0.019 | 9F9FFF |
| 0.750 | | V | MRPS27 | 0.616 | B3B3FF | 0.835 | 2323FF | 0.725 | 0.155 | 6B6BFF |
| 0.775 | | I | MRPS17 | 0.646 | 9F9FFF | 0.651 | 9C9CFF | 0.648 | 0.003 | 9D9DFF |
| 0.800 | | C | MRPS24 | 0.768 | 4F4FFF | 0.426 | FFCFCF | 0.597 | 0.242 | BFBFFF |
| 0.825 | | H | MRPS16 | 0.758 | 5656FF | 0.616 | B3B3FF | 0.687 | 0.101 | 8484FF |
| 0.850 | | Z | MRPS34 | 0.660 | 9696FF | 0.633 | A8A8FF | 0.646 | 0.019 | 9F9FFF |
| | | U | MRPS26 | 0.763 | 5252FF | 0.633 | A8A8FF | 0.698 | 0.092 | 7D7DFF |

**Table S10:** **Fold change values obtained from proteomic analysis used to color specific subunits on the cryo-EM structure of the mitoribosomal SSU. Red indicates strongly reduced protein abundance; blue indicates weakly reduced protein abundance.**
